# Prospective study of *Candida auris* nucleic-acids in wastewater solids in 190 wastewater treatment plants in the United States suggests widespread occurrence

**DOI:** 10.1101/2024.03.25.24304865

**Authors:** Alessandro Zulli, Elana M. G. Chan, Bridgette Shelden, Dorothea Duong, Xiang-Ru S. Xu, Bradley J. White, Marlene K. Wolfe, Alexandria B. Boehm

## Abstract

*Candida auris* is an emerging, multidrug-resistant fungal pathogen that poses a significant public health threat in healthcare settings. Despite yearly clinical cases rapidly increasing from 77 to 8,131 in the last decade, surveillance data on its distribution and prevalence remains limited. We implemented a novel assay for *C. auris* detection on a nationwide scale prospectively from September 2023 to March 2024, analyzing a total of 13,842 samples from 190 wastewater treatment plants across 41 U.S. states. Assays were extensively validated through comparison to other known assays and internal controls. Of these 190 wastewater treatment plants, *C. auris* was detected in the wastewater solids of 65 of them (34.2%) with 1.45% of all samples having detectable levels of *C. auris*. Detections varied seasonally, with 2.00% of samples positive in autumn versus 1.01% in winter (p<0.0001). The frequency of detection in wastewater was significantly associated with states having older populations (p<0.001), sewersheds containing more hospitals (p<0.0001), and sewersheds containing more nursing homes (p<0.001). These associations are in agreement with known *C. auris* epidemiology. This nationwide study demonstrates the viability of wastewater surveillance for *C. auris* surveillance, and further highlights the value of wastewater surveillance when clinical testing is constrained.

## Introduction

Healthcare associated infections are rapidly emerging as a global public health threat due to increasing levels of antimicrobial resistance and an aging worldwide population (1–3). Among these infections, *Candida auris,* first identified in 2009 in Japan, is an opportunistic, drug-resistant fungal pathogen with significant levels of associated morbidity and mortality (3–6). It has now spread to over 40 countries and has been designated as a priority pathogen by both international organizations, such as the World Health Organization (WHO), and national organizations, such as the Center for Disease Control and Prevention (CDC) in the United States, where it is now a nationally notifiable condition (4, 6, 7). The estimated crude mortality rate within the United States for *C. auris* infections was 34% between 2017-2022 (5).

*C. auris* prevention and control efforts are exacerbated by the fungus’ hardiness on fomites and high levels of transmissibility (6–8). The fungus has been found to persist for at least 7 days on both moist and dry surfaces, increasing the frequency and chance of transmission events in healthcare settings (9, 10). Of particular concern are high-touch plastic surfaces within healthcare facilities where *C. auris* was found to be viable for up to 14 days (10). In addition, *C. auris* is resistant to many common disinfectants so proper decontamination protocols are difficult and time intensive in already strained healthcare facilities (11). Further complicating containment efforts are the different presentations in colonized individuals compared to those with acute infections, with most colonized individuals presenting as asymptomatic (4). A lack of screening in high-risk patients and misidentification of specimens as other *Candida* subspecies has also contributed to the rapid spread of *C. auris* (5, 12). In the United States, clinical cases rose from 77 between 2013-2016 to 8,131 in 2022 and are projected to continue rising (5, 13).

Despite this tremendous increase in cases and the accompanying screening efforts, clinically available data are still sparse, with many institutions not speciating *Candida* cases resulting in underreporting cases in long-term care facilities and nursing homes (13, 14). Many of these facilities do not have the necessary equipment or human capital to implement speciation efforts and screening, which has been shown to be a necessary part of successful containment efforts (12). Alternative approaches to clinical surveillance are therefore necessary to better track both the spread and severity of outbreaks. Wastewater represents a naturally composite sample which is particularly useful for disease surveillance as it contains urine, feces, vomit, saliva, sloughed skin cells, sputum, and other excretions. Wastewater surveillance has been successfully used to track a wide variety of human pathogens including enterovirus, SARS-CoV-2, influenza A virus, and flavivirus (15–17). Like these viruses, *C. auris* has been detected in urine and stool samples from colonized or infected individuals, though it is typically diagnosed through presence in the blood or dermis of infected patients (4, 10, 18). Furthermore, recent studies have demonstrated the persistence and subsequent detection of *C. auris* in wastewater samples in Nevada and Florida (19). Early detection of this emerging pathogen through environmental surveillance may complement clinical surveillance efforts and inform containment and screening procedures (20).

In this paper, we present the results of a United States-wide wastewater monitoring effort for *C. auris* and compare the data to publicly available, recent, clinical case data. We also compare results to available demographic indicators and healthcare facility locations. Detections of *C. auris* in wastewater demonstrate wide geographical occurrence, and we demonstrate that these wastewater detections appear to be associated with age and presence of hospitals and nursing homes in adjacent communities. These results demonstrate the advantages of using wastewater surveillance for the detection of emerging pathogens and for supplementing clinical testing efforts.

## Methods

### RT-PCR assays

We designed a novel probe for *Candida auris* to use in conjunction with previously published specific and sensitive forward and reverse primers (21). The assay targets the region of the genome encoding the 5.8S ribosomal RNA, all of ITS2, and a fragment of 28S ribosomal RNA. To design the probe, *C. auris* genome sequences were downloaded from the National Center for Biotechnology Information (NCBI) and aligned to identify conserved regions located between the primers. The probe was designed *in silico* using Primer3Plus (https://primer3plus.com/). Parameters used in the design process (e.g., sequence length, GC content, and melt temperatures) are provided in Table S1. The primers and probe were then screened for specificity *in silico* by blasting the sequences in NCBI and *in vitro* against fungal panels, or cDNA sequences. The *in silico* analysis was performed by excluding all *C. auris* genomes from a BLAST search, thereby only giving off-target matches (22).

The primers and probe were tested *in vitro* for specificity and sensitivity using a fungal panel (NATCTVPOS-BD, Zeptomatrix Buffalo, NY) and *Candida auris* Satoh et Makimura (ATCC MYA-5001) purchased from American Type Culture Collection (ATCC, Manassas, VA). The fungal panel NATCTVPOS-BD includes chemically inactivated *Candida albicans*, *C. krusei*, *C. glabrata*, and *Trichomoniasis vaginalis*. Nucleic acids were extracted from fungi using Chemagic Viral DNA/RNA 300 Kit H96 for Chemagic 360 (PerkinElmer, Waltham, MA).

Nucleic acids were used undiluted as template in digital droplet RT-PCR (ddRT-PCR) singleton assays for sensitivity and specificity testing in single wells. The concentration of targets used in the *in vitro* specificity testing was between 10^3^ and 10^4^ copies per well. Negative RT-PCR controls were included on each plate.

We used an RT-PCR assay for the in vitro sensitivity and specificity testing as the goal was to ultimately multiplex the *C. auris* assay in a panel that includes RNA-genome viruses. The implications of the use of RT-PCR, and a comparison to results obtained using PCR are further investigated, as described below. Regardless, this means the assay detects both DNA and RNA targets.

### Clinical surveillance data

The National Notifiable Disease Surveillance System (NNDSS) is a nationwide collaboration to which all levels of healthcare providers share health information about nationally notifiable infectious and noninfectious diseases (23). Case reports are compiled on a weekly basis using uniform case definitions from 50 states, the District of Columbia, New York City, and 5 territories (American Samoa, Guam, Northern Mariana Islands, Puerto Rico, U.S. Virgin Islands) (23). For this study, we used publicly available information on NNDSS for *Candida auris* from 11 September 2023 to 1 March 2024 (https://data.cdc.gov/api/views/x9gk-5huc/rows.csv?accessType=DOWNLOAD). Dates of case determination varied but included the following in order of preference: date of disease start, date of diagnosis, date of laboratory result, date of first report to public health system, or date of state report (24). States which did not report clinical case data were omitted from analyses. It should be noted that this database is known to be incomplete as many local jurisdictions do not submit data; additional data on C. auris cases or Candida cases may be available from other local or regional sources that are not publicly available or currently updated.

### Wastewater data: sample collection

Wastewater measurements were made prospectively as part of a wastewater surveillance program. Between 11 September 2023 and 1 March 2024, wastewater samples (either 24-hour composited influent or grab samples from the primary clarifier, Table S1) were collected by wastewater treatment plant (WWTP) staff using sterile containers. Samples were typically obtained approximately three times per week and shipped overnight to the laboratory at 4°C where they were processed immediately with no storage. Samples were collected from 190 distinct WWTPs across a total of 41 states over the course of the study period (Figure 1). A total of 13,842 samples were collected and analyzed as part of this study.

**Figure 1.**
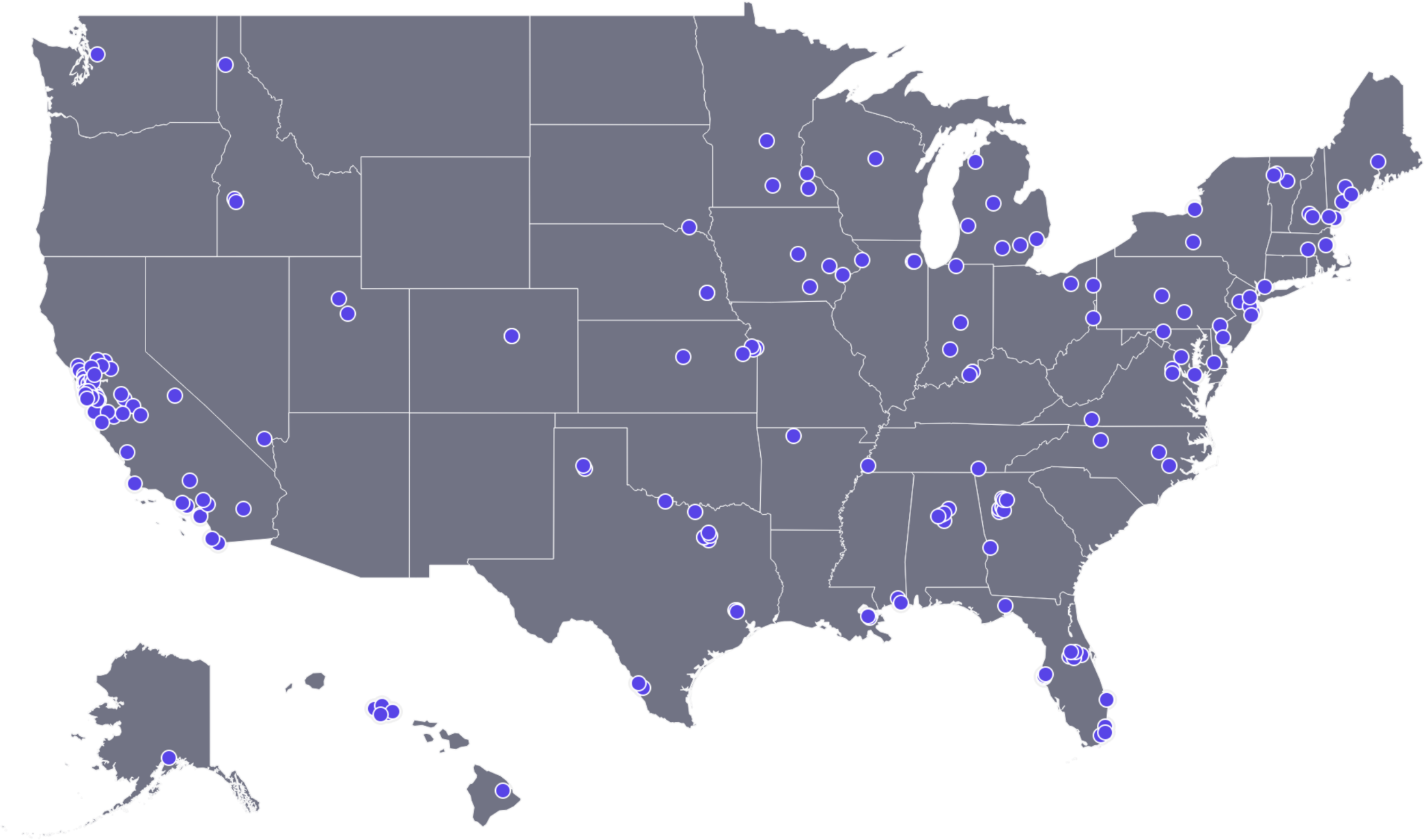
Location of wastewater treatment plants (WWTPs) with *Candida auris* DNA data included in the paper. Each blue dot represents the location of a WWTP.

### Wastewater data: pre-analytical processing

*C. auris* is expected to be associated with the solid phase of wastewater based on previous empirical measurements across liquid and solid phases in wastewater and its size (diameter = 2.5-5 µm), so in this study we made measurements in the solid phase of wastewater (25, 26). Details of the isolation of solids from the samples are provided in other peer-reviewed publications (27). In short, samples were centrifuged to dewater the solids. One aliquot of dewatered solids was used for nucleic-acid extractions and another was used to determine the dry weight of the solids using an oven (27). Approximately 75 mg of dewatered solids was added per milliliter of DNA/RNA shield (Zymo, Irvine, CA) spiked with bovine coronavirus vaccine as a positive extraction control. This mixture of solids and buffer was previously shown to have minimal inhibition.(28) The mixture was homogenized after the addition of grinding balls, and centrifuged; the supernatant was used immediately for nucleic-acid extractions. Nucleic-acids were extracted from 6-10 replicate aliquots of the supernatant using the Chemagic Viral DNA/RNA 300 kit H96 for the Perkin Elmer Chemagic 360 (Perkin Elmer, Waltham, MA) followed by PCR inhibitor removal with the Zymo OneStep-96 PCR Inhibitor Removal kit (Zymo Research, Irvine, CA). Whether 6 or 10 replicates were used is specified in Table S1. The suspension volume entered into the nucleic-acid extraction process was 300 µl and 50 µl of nucleic-acids were retrieved after the inhibitor removal kit. Negative extraction controls consisted of BCoV-vaccine spiked DNA/RNA Shield. Nucleic-acids were used immediately as template in PCR as described next, with no storage.

### Wastewater data: analytical processing

We used droplet digital reverse transcription polymerase chain reaction (ddRT-PCR) to measure nucleic-acids in wastewater. We measured pepper mild mottle virus (PMMoV) as an endogenous positive control as concentrations tend to be extremely elevated in wastewater samples; we measured BCoV as an endogenous, spiked-in control (28–30). Methods applied to wastewater solids to measure PMMoV and BCoV in a duplex reaction are provided in detail elsewhere (27). The *C. auris* assay (Table 1) was run in multiplex using a probe-mixing approach. The exact assays that *C. auris* was multiplexed with varied slightly over the duration of the project for some of the plants ( information is provided in Table S2). Multiplexing *C. auris* with these targets did not affect *C. auris* quantification (see SI Figure S1). The results from those other assays are not provided herein. The PMMoV/BCoV duplex and *C. auris* multiplex assays were run on 96-well plates. Each 96-well PCR plate of wastewater samples included PCR positive controls for each target assayed on the plate in 1 well, PCR negative no template controls in two wells, and extraction negative controls in two wells. PCR positive controls consisted of *C. auris* DNA. Each of the 6 or 10 replicate nucleic-acid extracts were run in their own wells to measure *C. auris*. Two randomly selected or 10 replicate nucleic-acid extracts were run in their own wells to measure PMMoV and BCoV (Table S1).

**Table 1.**
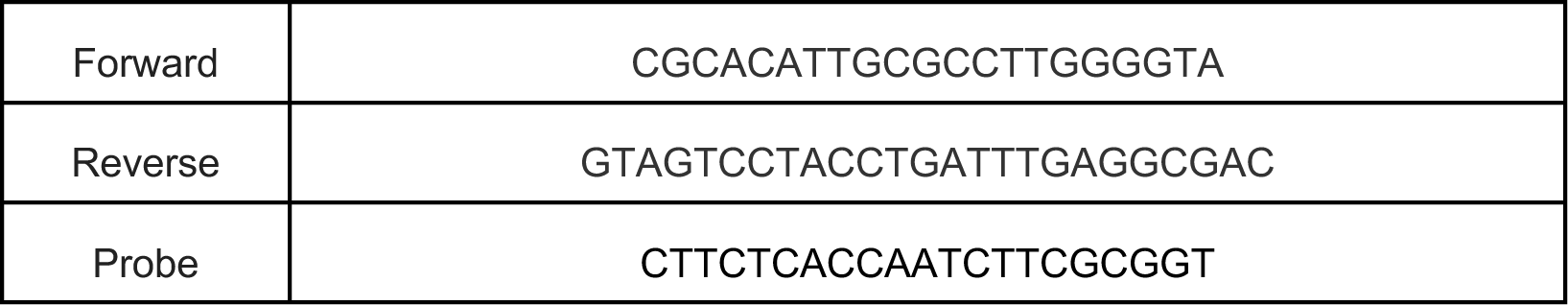
Primers and probes used in this study for detection of *C. auris* nucleic acids. Primers and probes were purchased from Integrated DNA Technologies (Coralville, IA, USA). The probes contained fluorescent molecule HEX and quenchers (5′ HEX/ZEN/3′ IBFQ); HEX, hexachloro-fluorescein; ZEN, a proprietary internal quencher from Integrated DNA Technologies (Coralville, IA, USA); and IBFQ, Iowa Black FQ. Amplicon size is 163 basepairs.

ddRT-PCR was performed on 20-µl samples from a 22 µl-reaction volume, prepared using 5.5 µl of template mixed with 5.5 µl of One-Step RT-ddPCR Advanced Kit for Probes (Bio-Rad 1863021), 2.2 µl of 200 U/µl Reverse Transcriptase, 1.1 µl of 300 mM dithiothreitol (DTT), and primers and probes mixtures at a final concentration of 900 nM and 250 nM, respectively. Primer and probes for assays were purchased from Integrated DNA Technologies (IDT, San Diego, CA) (Table 1). *C. auris* was measured in reactions with undiluted template whereas PMMoV and BCoV were run on template diluted 1:100 in molecular grade water. It is important to note that an RT step was used because during the prospective study, we were measuring a large number of RNA genome viruses. Therefore, the *C. auris* target quantified should be interpreted as copies of RNA plus DNA.

Droplets were generated using the AutoDG Automated Droplet Generator (Bio-Rad, Hercules, CA). PCR was performed using Mastercycler Pro (Eppendforf, Enfield, CT) with with the following cycling conditions: reverse transcription at 50°C for 60 minutes, enzyme activation at 95°C for 5 minutes, 40 cycles of denaturation at 95°C for 30 seconds and annealing and extension at 59°C (for *C. auris*) or 56°C (for PMMoV and BCoV) for 30 seconds, enzyme deactivation at 98°C for 10 minutes then an indefinite hold at 4°C. The ramp rate for temperature changes were set to 2°C/second and the final hold at 4°C was performed for a minimum of 30 minutes to allow the droplets to stabilize. Droplets were analyzed using the QX200 or the QX600 Droplet Reader (Bio-Rad). A well had to have over 10,000 droplets for inclusion in the analysis. All liquid transfers were performed using the Agilent Bravo (Agilent Technologies, Santa Clara, CA).

Thresholding was done using QuantaSoft™ Analysis Pro Software (Bio-Rad, version 1.0.596) and QX Manager Software (Bio-Rad, version 2.0). Replicate wells were merged for analysis of each sample. In order for a sample to be recorded as positive, it had to have at least 3 positive droplets. Concentrations of RNA targets were converted to concentrations in units of copies (cp)/g dry weight using dimensional analysis. The total error is reported as standard deviations. Three positive droplets across 6 merged wells corresponds to a concentration between ∼1000 cp/g; the range in values is a result of the range in the equivalent mass of dry solids added to the wells. Data collected as part of the study are available from the Stanford Digital Repository (https://purl.stanford.edu/hv291tt0888).

### Wastewater: Confirmatory analyses

We selected seven samples (Table S3) with relatively high concentrations of *C. auris,* as measured above, to test another published *C. auris* assay by Barber et al. (25) for detecting *C. auris* in wastewater. The assay used by Barber et al. targets the same region of the *C. auris* genome as the assay in Table 1 but uses different primers and probes (Table S4) (25). Archived nucleic acid extracts, stored at −80°C, from those samples were thawed and then run as template using the Barber et al. assay as well as the assay described herein (Table 1). The samples were run in duplex with an assay that targets the N gene of SARS-CoV-2 (28). *C. auris* was run using a probe labeled with fluorescein amidite (FAM). The methods described above for the analytical procedures (number of replicate wells, negative and positive controls) were used.

These same seven samples were assayed with and without the reverse transcription step. Nucleic-acid extracts were thawed and run exactly as described for the prospective study except that the RT and DTT were not added to the mastermix.

We selected a subset of five of the seven samples (Table S3) for Sanger sequencing of the PCR amplicon. One µl of archived (at −80°C) nucleic acid extracts was used as the template in a 50-µl PCR reaction prepared according to manufacturer’s protocol with Q5® High-Fidelity 2X Master Mix (New England Biolabs, Ipswich, MA) using the primers in Table 2. The thermocycler program was run at an annealing temperature of 67°C for 30 seconds and extension at 72°C for 30 seconds. *C. auris* DNA was used as a positive control. PCR products were run on a 2% agarose gel stained with SYBR Safe gel stain (Thermofisher, Fremont, CA) and amplicons of the expected size (259 bp) were excised and gel purified using Zymo gel DNA recovery kit (Irvine, CA). The purified DNA was then sent to Molecular Cloning Laboratories (South San Francisco, CA) for sanger sequencing. Sequences were downloaded and aligned with *C. auris* genomes using Snapgene software.

### Demographic data

*C. auris* hospitalizations are most common among older adults (5). We used 5-year estimates from the 2022 American Community Survey (ACS) at the state level to identify the median age of states with participating WWTPs (n = 41 states) (31).

### Hospitals and nursing homes data

*C. auris* is particularly a concern in hospitals and nursing homes (5, 14, 20). We obtained the locations of hospitals and nursing homes in the United States from the US Department of Homeland Security’s Homeland Infrastructure Foundation-Level Data (HIFLD) database (https://gii.dhs.gov/HIFLD) (32, 33). We selected all types of hospitals and nursing homes with an “open” status and then determined the number of hospitals and nursing homes in each sewershed using the Tabulate Intersection geoprocessing tool in ArcGIS Pro (version 3.1.1). Sewershed boundaries were provided by most WWTPs (n = 115).

For WWTPs that did not provide a sewershed (n = 75), we approximated sewershed boundaries based on the zip codes serviced by the WWTP and USA ZIP Code Boundaries geospatial data set published by Esri (Source: TomTom, US Postal Service, Esri) (34).

### Data analysis

We made comparisons on the basis of percent of positive detections in wastewater. Percentage of positive detections of *C. auris* was calculated by counting all positive observations for a single WWTP and dividing by the total number of observations. Wastewater measurements in each state were aggregated by Morbidity and Mortality Weekly Report (MMWR) week, and we calculated the percentage of wastewater measurements that were positive for *C. auris* each week.

Spearman’s rank correlation coefficients were used to assess the association between weekly (MMWR weeks) publicly available NNDSS clinical data and wastewater percent positive detections (hereafter “detections”) of *C. auris* for each state (n=18 states with wastewater and clinical data). Case data from NNDSS, available on a weekly basis for each state, was used after adjusting for population (cases per million people, Fig 2a).

**Figure 2:**
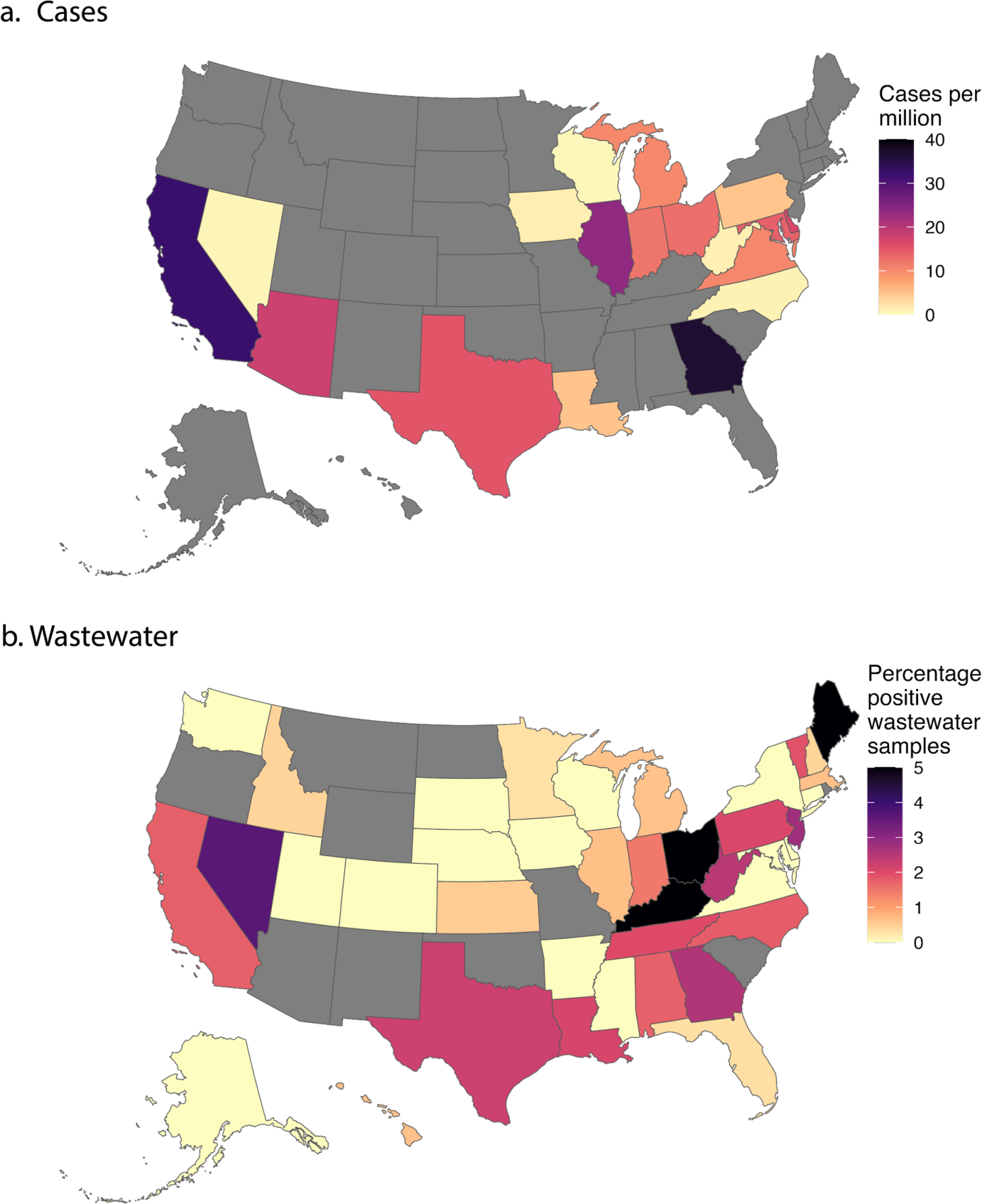
Map showing percentage of positive wastewater samples and population normalized case numbers from NNDSS across the United States. Figure 2a shows the normalized number of cases per million in each state as reported to NNDSS during the study period. Figure 2b shows the percentage of positive wastewater samples in each of the states surveilled during the study period. Yellow represents low normalized values while black represents high normalized values. Gray states represent areas without available data.

We tested whether concentrations measured by the assay presented in Table 1 were different from those measured by the Barber et al. assay using RT-ddPCR on 9 representative samples (Table S3, Figure S2) (25). We ran the same samples using RT-ddPCR and ddPCR with no RT step (Table S3, Figure S2). Differences between these controls were assessed using a paired t-test.

We also assessed the relationship between *C. auris* percent positive detections and several other contextual and demographic factors. We explored seasonality by assessing *C. auris* detections in wastewater samples collected in fall (defined as Sept 1 to Nov 30) and winter (defined as Dec 1 to Feb 29). We then assessed the association between *C. auris* detections and age at the state level. We grouped WWTPs by those in states with a median age less than versus greater than or equal to the national median age (39 years) (31). Lastly, we assessed the association between *C. auris* detections and hospitals and nursing homes at the sewershed level. We grouped WWTPs by those with a sewershed intersecting (1) less than versus greater than or equal to the median number of hospitals among all sewersheds (median: 1 hospital) and (2) less than versus greater than or equal to the median number of nursing homes among all sewersheds (median: 7 nursing homes). For seasonality, we compared the percentage of wastewater samples positive for *C. auris* in each season. For age, hospitals, and nursing homes, we compared the percentage of wastewater samples that were positive for *C. auris* across WWTPs in the less than median group versus the greater than or equal to median group. The null hypothesis tested for each of these variables was that there were no differences between the two groups.

Statistical significance and 95% confidence intervals between the groups described above were assessed through bootstrapping (35, 36). For each variable and group, a distribution of values was generated through random sampling with replacement 10,000 times. Significance was then calculated using the proportion of values below our calculated statistic (i.e. whether the calculated statistics were above the 95% confidence intervals). This same method was used when assessing differences between assays (Fig S2) and seasonal differences in detection. In those cases, grouping was done based on assay and season, respectively.

## Results

### QA/QC

The *C. auris* assay was found to be sensitive and specific. The *in silico* analysis indicated no primer or probe sequence was more than an 85% match to off-target matches, and there was no overlap found between combinations of forward primer off-targets, reverse primer off-targets, and probe off-targets. These factors indicate a high level of specificity in the primer-probe combination used. The in vitro testing confirmed sensitivity and specificity of the *C. auris* assay as *C. auris* nucleic-acids were positive and the remaining non-target nucleic-acids were negative.

Results are reported following the Environmental Microbiology Minimal Information (EMMI) guidelines (see SI and Figure S3).(37) All positive and negative controls performed as expected, indicating acceptable assay performance. Median (IQR) BCoV recoveries across all wastewater samples were 1.08 (0.82, 1.43) indicating good recovery across all samples. Recoveries exceeding 1 are the result of uncertainties in the measurement of BCoV added to the buffer matrix. PMMoV levels were elevated in all samples indicating lack of gross extraction failures (median = 4.8×10^8^ cp/g, min = 5.0×10^5^ cp/g, max = 2.02×10^11^ cp/g).

As a further validation step, *C. auris* was quantified in a subset of samples also using the Barber et al. assay (Figure S2) (25). Concentrations measured by the two assays closely matched, with no statistically significant differences (p>0.05). Comparisons between concentrations measured using RT-ddPCR and ddPCR showed similar results (Figure S2); no significant differences were found (p>0.05). Additionally, the PCR amplicons obtained using the forward and reverse primers were sequenced and the sequences matched the *C. auris* genome segment targeted (Figure S4).

### National Overview

The study period spanned 11 September 2023 to 1 March 2024. Wastewater data were available from 190 distinct WWTPs in 41 states (Table S1). The number of WWTPs in these states ranged between 1–57 WWTPs (median: 2 WWTPs), with population coverage of the sewersheds ranging from 0.13% to 59.5% of the population of each individual state (median: 5.75%). The number of wastewater samples collected and analyzed from individual WWTPs ranged between 23–173 samples (median: 72 samples) during the study period. In total, 13,842 wastewater samples were collected and analyzed across all WWTPs during the study period. *C. auris* concentrations in these samples ranged from below the limit of detection to 1,456,751 cp/g, with 1.45% (n = 200 samples) of samples above the lowest detectable concentration. Sixty five (34.21%) WWTPs across 26 states had at least one detectable *C. auris* concentration during the study period, and the percentage of positive samples from these WWTPs ranged between 0.58%–46.48% (median: 1.49%). Time series heatmaps of *C. auris* detections at each of the 190 WWTPs are provided in Figure S5. Seasonally, 2.00% (n=6754) of observations were positive during autumn (defined as Sep 1 to Nov 30), and 1.01%(n=7245) of observations were positive during winter (defined as Dec 1 to Feb 29). This difference was statistically significant when assessed by a bootstrapping approach (p<0.0001).

Clinical data from the National Notifiable Disease Surveillance System (NNDSS) were downloaded for comparison to wastewater data, and no significant correlations were found between wastewater detections and population adjusted clinical case rates (Spearman’s rho between −0.10 and 0.23 for the 18 states, all p>0.05). Several states had obvious divergence between wastewater positivity rates and population adjusted NNDSS clinical case rates (**Figure 2)**. For example, in Maine, 13.3% of wastewater samples were positive for *C. auris* despite not reporting any clinical cases during the study period to NNDSS (**Figure 2**). Twelve states which did not report *C. auris* cases to the NNDSS were found to have positive detections of *C. auris*. **Figure 3** shows percentage positive detections as a function of population normalized clinical cases and highlights the limited clinical data available.

**Figure 3.**
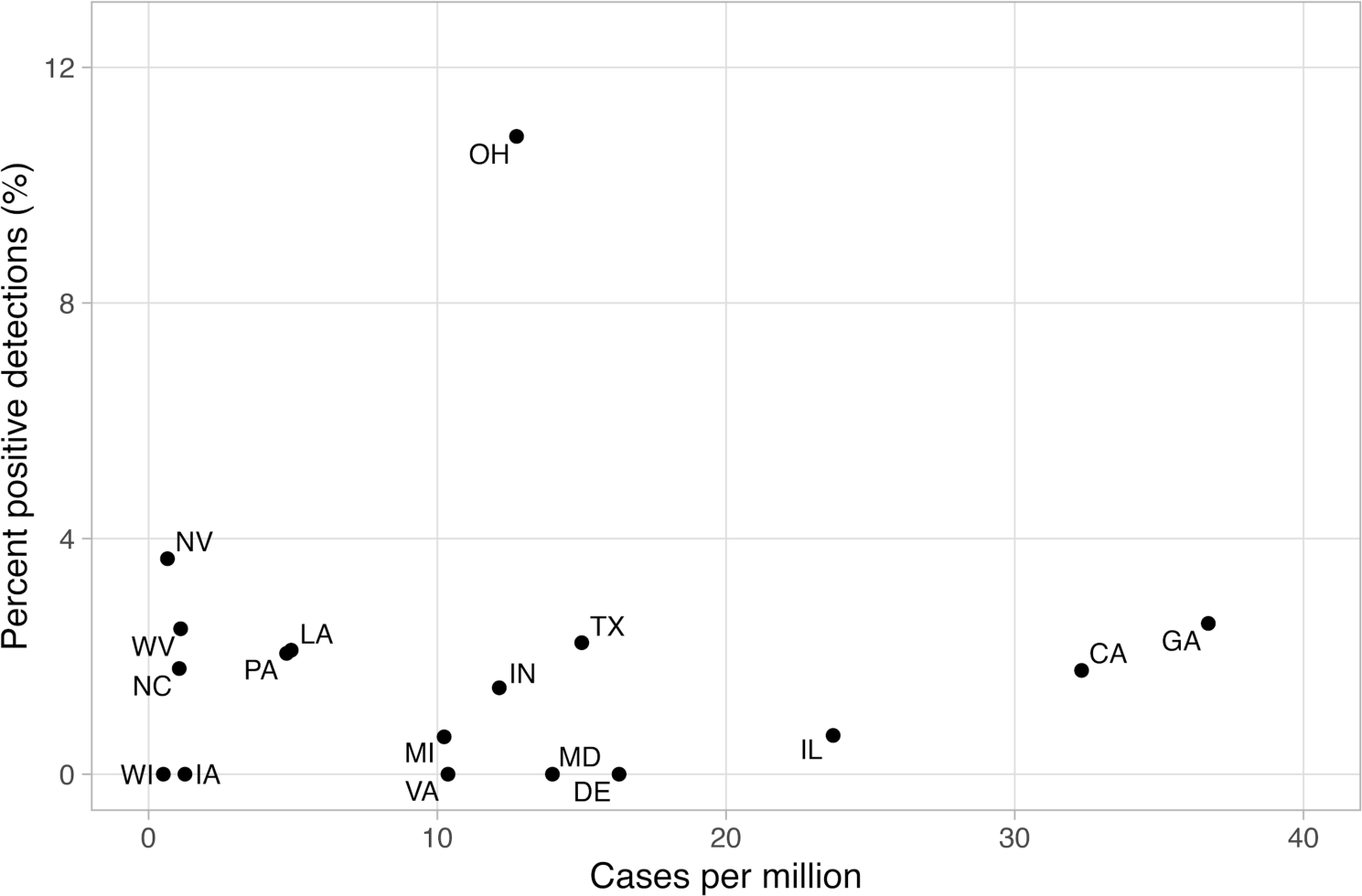
Comparison of wastewater percent positive detections and total clinical cases as reported by NNDSS. Only states with clinical data reported to NNDSS are included. Each labeled point represents the percentage of positive wastewater samples in that state and the number of cases reported to NNDSS. The state abbreviation is above each data point.

## Population demographics and healthcare facility locations and *Candida auris*

WWTPs were grouped using three separate statistics: median age of the state, number of hospitals in the sewershed, and number of nursing homes in the sewershed (Fig 4). The median age in the United States is 39 years as of the latest American Community Survey (ACS) data (31). Seventy-eight WWTPs were located in a state with a median age at or above the national median age; 112 WWTPs were located in a state with a median age below the national median age. As shown in **Figure 4a**, among WWTPs in states with a median age at or above the national median age, 2.03% (95% CI: [0.80-3.50]) of all wastewater samples were positive for *C. auris*. Among WWTPs in states with a median age below the national median age, 0.83% (95% CI: [0.34-1.38]) of all wastewater samples were positive for *C. auris*. This difference was statistically significant when assessed by bootstrapping (p < 0.001).

**Figure 4.**
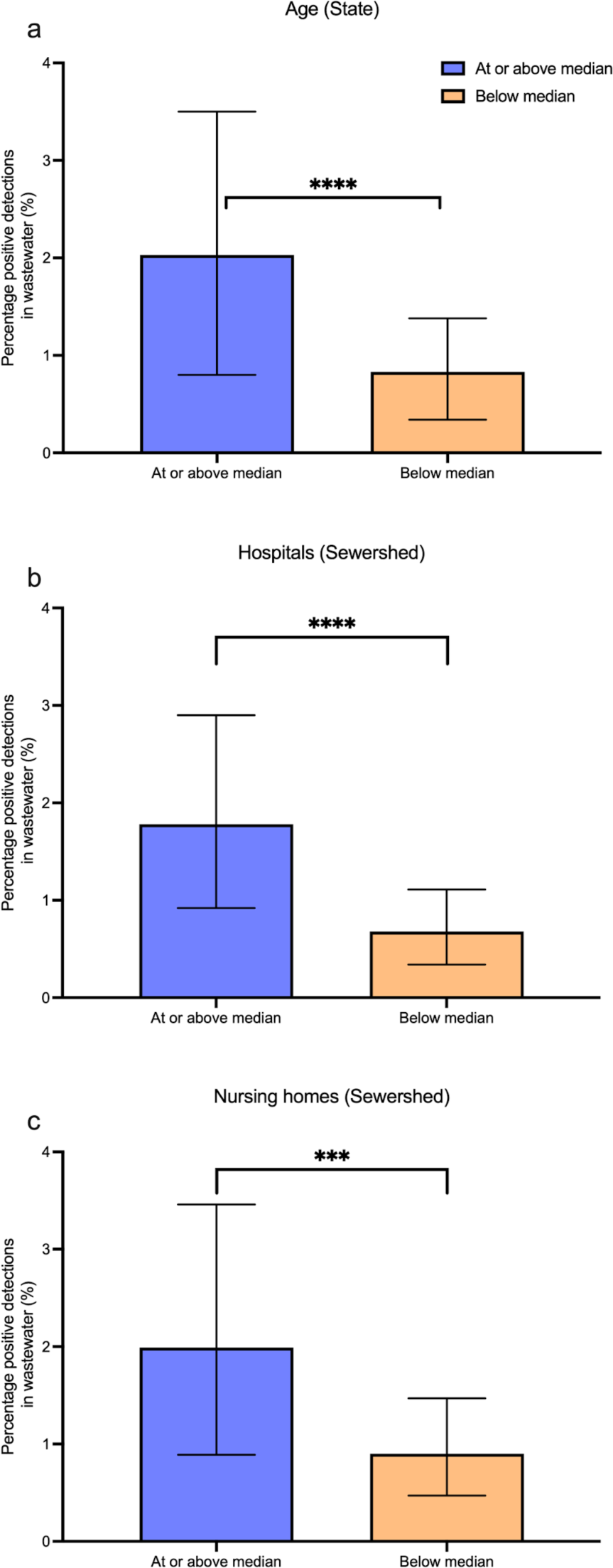
Comparison of wastewater detections across different groups. *** indicates p<0.001, **** indicates p<0.0001. Figure 4a shows the percentage of positive samples for states at or above the median age compared to those below. Figure 4b shows the percentage of positive samples when grouped by number of hospitals at the sewershed level. Figure 4c shows the same statistic when grouped by number of nursing homes, again at the sewershed level. Error bars represent 95% confidence intervals derived from the bootstrapping approach.

The median number of hospitals in the sewersheds was 1 hospital (range: 0–74 hospitals). WWTPs were grouped by whether they contained 0 hospitals (fewer than the median, n = 54) or 1 or more hospitals in their sewersheds (equal to or greater than the median, n = 136). We found that 1.78% (95% CI: [0.92-2.90], n = 10,019) of samples from WWTPs with at least 1 hospital in their sewersheds were positive for *C. auris* whereas 0.62% (95% CI: [0.34-1.11], n = 3,819) of samples from WWTPs with no hospital in their sewershed were positive for *C. auris* (**Figure 4b**). This difference was significant when assessed by bootstrapping (p < 0.0001).

The median number of nursing homes in the 190 sewersheds was 7 (range: 0–420 nursing homes). WWTPs were grouped by whether they contained 0–6 nursing homes (fewer than the median, n = 91) or 7 or more nursing homes in their sewersheds (equal to or greater than the median, n = 99). We found that 1.92% (95% CI: [0.89–3.46], n = 7,544) of samples from WWTPs with 7 or more nursing homes in their sewersheds were positive for *C. auris* whereas 0.87% (95% CI: [0.47–1.47], n = 6,294) of samples from WWTPs with 0–7 nursing homes in their sewersheds were positive for *C. auris* (**Figure 4c**). This difference was significant when assessed by bootstrapping (p < 0.001).

## Discussion

Previous studies at the sewershed scale have shown that detection of *C. auris* in wastewater is indicative of ongoing outbreaks in the contributing population (19, 25). To our knowledge, this study represents the first nationwide wastewater monitoring effort of *C. auris* and comparison to national clinical case data (23). We first demonstrate the sensitivity and specificity of the assay used in this study through comparisons to published assays, sequencing (Figure S2, S4), and in silico analyses. The widespread detection of *C. auris* in wastewater suggests a significant gap in clinical case data reported to the NNDSS. Indeed, it is known that many local jurisdictions do not provide data for inclusion in NNDSS. We found that *C. auris* detections are more frequent in states with older populations and in sewersheds with higher numbers of nursing homes and hospitals. These findings are consistent with *C. auris* infections being more common in older, more susceptible populations and further support the assertion that clinical case data may be incomplete.

*C. auris* has gone from initial detection in the United States in 2013 to being declared an “Urgent” and notifiable pathogen in 2019 by the United States (CDC) (14, 20, 38). The rate of testing volume and clinical positives has rapidly increased, with 17 states identifying their first *C. auris* cases between 2019 to 2021 (14). Despite this, clinical testing for *C. auris* is far from ubiquitous and not conducted uniformly across the United States (14, 39). On-site PCR testing of clinical specimens is rare, and samples often have to be sent to public health or reference laboratories, further increasing the cost of testing and the turnaround time before the data reaches public health officials (39). Detections of *C. auris* nucleic-acids in wastewater collected throughout the United States demonstrate a need for increased clinical testing and reporting capacity, and the capability of wastewater testing to serve as a sentinel surveillance mechanism for emerging pathogens.

*C. auris* is an opportunistic pathogen that often infects the elderly and immunocompromised within long-term healthcare facilities (10, 20, 40). Following our detection of *C. auris*, we investigated potential demographics and healthcare facilities related to its pathology using data from the ACS and US Department of Homeland Security’s HIFLD database (31, 41). We analyzed age, number of hospitals, and number of nursing homes as plausible factors impacting the presence of *C. auris* within a sewershed or state. We found significant differences between the percentage of positive detections of *C. auris* in states with a median age less than versus greater than or equal to the national median age. Similarly, we found significant differences between the percentage of positive detections of *C. auris* in sewersheds with less than versus greater than or equal to the median number of hospitals and nursing homes. States with a population above the national median age were more than three times as likely to detect *C. auris* in a given wastewater sample. At the sewershed level, locations with 1 or more hospitals had three times the amount of *C. auris* detections than locations with no hospitals. Similarly, sewersheds with greater than or equal to the median number of nursing homes had over twice as many detections of *C. auris* as those with fewer than the median number of nursing homes. These findings are consistent with the known epidemiology of *C. auris*, as infected patients’ median age is 68 and outbreaks are prevalent in long-term healthcare facilities and nursing homes (5, 14). The presence of an at-risk population appears to be strongly associated with *C. auris* wastewater detections, suggesting public health efforts are appropriately focused on these at-risk populations.

Significant steps were taken to ensure the validity of detection results. A similar assay was used by Barber et al. for the detection of *C. auris* in southern Nevada during an outbreak (25). We tested nine samples from three different states using both the assay presented in this study and the one used by Barber et al. (25). No statistically significant difference was found between the two assays, which both targeted the same region of an internally transcribed spacer region. Our assay was further validated using a subset of 5 of these 9 samples through sequencing. The amplicons generated by our forward and reverse primers independently and accurately matched the reference amplicon, indicating specific amplification (Figure S4). Together, these results demonstrate that the assay used in this study is highly specific. Finally, we used an RT-PCR assay to detect *C. auris* which means the assay should detect both RNA and DNA. We measured some samples using both RT-PCR and PCR and the results indicated that most of the target nucleic acids in those samples were DNA. This was somewhat surprising since we expect the DNA target (located in the region of the genome that encodes ribosomal RNA) should be transcripted into RNA by *C. auris*. Future work will need to be done to further investigate the relative proportion of target RNA versus DNA *C. auris* targets in wastewater. Regardless, the use of an RT-PCR assay, allowing detection of both RNA and DNA, should increase the sensitivity of the assay (by detecting both nucleic-acids) yet perhaps complicate interpretation of measured concentrations. For this reason, we only use the data in presence/absence format.

Overall, our results demonstrate that *C. auris* wastewater surveillance can provide timely information on geographical distribution and help identify at-risk populations; wastewater data can be available within 24-48 hours of sample collection. As an emerging disease, clinical testing for *C. auris* largely depends on shipping samples to public health laboratories (7, 13, 14). Wastewater surveillance can help fill this gap for municipalities and states without the capital to fund testing systems, providing community level information as to the prevalence, and more importantly, the spread of *C. auris*. These data are critical to public health responses, particularly when dealing with pathogens that do not or did not have robust clinical testing systems such as Dengue virus, SARS-CoV-2, and *C. auris* (14, 16, 17, 39). *C. auris* wastewater surveillance data can then be used to implement stricter screening protocols, cleaning protocols, and other non-pharmaceutical interventions that can prevent the spread of illness amongst vulnerable populations (9, 10, 20).

There are limitations associated with this study. Importantly, the availability of clinical data was limited due to the recent classification in 2018 of *C. auris* as a notifiable disease, making in-depth analyses of associations between wastewater detections and clinical data on infections unfeasible. In addition, it is well understood that the NNDSS database may not contain all the information on C. auris positive cases in the United States; local jurisdictions may have local data to inform public health response to infections and those data were not available for this work. Local jurisdictions are encouraged to use these wastewater data to compare to their local case rates. Further, wastewater sampling was not uniform across the United States or within individual states, which could introduce biases into the analyses. Within the demographic data available at the sewershed level, information was limited as to the size of nursing homes and hospitals, which might be important factors controlling associations with wastewater data on *C. auris.* Lastly, we were unable to link specific wastewater concentrations to population-level incidence. Further experiments are necessary to understand the shedding patterns of *C. auris* in human excretions as to provide this direct link to disease occurrence in the contributing population. These experiments are particularly important to the determination of the significance of wastewater detections, as *C. auris* contributions to wastewater could potentially be from colonized individuals or individuals with acute infection. Finally, an additional limitation is that we cannot rule out the potential for *C. auris* detections in wastewater to be from zoonotic or environmental sources. Detections have been shown to match underlying expected demographic indicators such as age and presence of nursing homes, further lending credence to the source of detections being human. While there is no evidence to support the possibility of zoonotic or environmental contributions of *C. auris* nucleic-acids to wastewater, further research is needed to see if such evidence might exist. There have been case reports of *C. auris* infections in canines, and *Candida spp*. have been detected using culture and other molecular methods from biofilms in sewer pipes (42–45). It should be noted that wastewater detection of Candida auris collected at the wastewater treatment plant scale does not imply community-spread of the pathogen, but is consistent with its presence in one or more individuals within the sewershed.

## Data Availability

All data produced are available online at the Stanford Digital Repository.

https://purl.stanford.edu/hv291tt0888

## Acknowledgements

We acknowledge all the wastewater treatment plant staff who provided samples for this project.

