## Supporting Information for "Prospective study of *Candida auris* nucleic-acids in wastewater solids in 190 wastewater treatment plants in the United States suggests widespread occurrence"

**Additional details related to EMMI guidelines.** One-hundred forty (140) wastewater samples from the study were selected at random for this analysis; this represents ~10% of the samples processed in the study. The randomly selected samples were from the samples for which 10 replicate wells were run for C. auris as these raw data were relatively more easily accessible. The average (standard deviation) number of partitions (droplets) for the across 10 replicate wells was 173,777 (29,518) for the reaction for *C. auris*. The volume of the partitions, as reported by the machine vendor is 0.00085 µL. The average (standard deviation) of copies per partition for the C. auris target is $1.37 \times 10^{-6}$ ($3.14 \times 10^{-6}$). An example fluorescent plot from the QX600 (6 color reader) is included in the Stanford Digital Repository with the deposited wastewater data (https://purl.stanford.edu/hv291tt0888).

**Table S1.** Plant characteristics. Plants marked with an asterisk (*) are plants where 10 replicate wells were used for digital droplet PCR (SCAN plants in caption of Table S2), the remaining plants had 6 wells used to measure C. auris, and 2 for PMMoV and BCoV. The site name, location (town, and state), state where plant is located, and whether the sample originally supplied to us was a 24 hour composite sample of liquid influent ("liquid") or a sample from the primary clarifier ("solids"). The population served is also provided.

| Site Name | Location | State | Sample Type | Population served |
|---|---|---|---|---|
| Valley Creek Water Reclamation Facility | Bessemer, AL | Alabama | Liquids | 225,000 |
| Cahaba River Water Reclamation Facility | Cahaba River, Birmingham, AL | Alabama | Liquids | 95,000 |
| Five Mile Creek Water Reclamation Facility | Fultondale, AL | Alabama | Liquids | 77,000 |
| Turkey Creek Water Reclamation Facility | Pinson, AL | Alabama | Liquids | 30,000 |
| Village Creek Water Reclamation Facility | Village Creek, Birmingham, AL | Alabama | Liquids | 200,000 |
| John M. Asplund Water Pollution Control Facility | Anchorage, AK | Alaska | Liquids | 220,000 |
| City of Harrison Wastewater Treatment Plant | Harrison, AR | Arkansas | Liquids | 15,000 |
| Coastal Treatment Plant | Coastal, Laguna Niguel, CA | California | Liquids | 48,000 |
| Central Contra Costa Sanitary District | Contra Costa County, CA | California | Solids | 484,800 |
| City of Davis Wastewater Treatment Plant | Davis, CA | California | Solids | 68,000 |
| Esparto Wastewater Treatment Facility | Esparto, CA | California | Liquids | 4,006 |
| Fairfield-Suisun Sewer District | Fairfield, CA | California | Solids | 155,000 |
| [Fremont Basin] - Raymond A. Boege Alvarado WWTP | Fremont, CA | California | Liquids | 229,476 |
| South County Regional Wastewater Authority * | Gilroy, CA | California | Solids | 110,338 |

| | | | | |
|---|---|---|---|---|
| Sewer Authority Mid-Coastside | Half Moon Bay, CA | California | Liquids | 28,000 |
| City of Hollister Domestic Water Recycling Facility | Hollister, CA | California | Liquids | 42,000 |
| Valley Sanitary District | Indio, CA | California | Solids | 91,765 |
| JB Latham Treatment Plant | JB Latham, Laguna Niguel, CA | California | Liquids | 120,000 |
| Lancaster Water Reclamation Plant | Lancaster, CA | California | Liquids | 200,000 |
| Las Gallinas Valley Sanitary District | Las Gallinas, San Rafael, CA | California | Liquids | 30,000 |
| Lompoc Regional Wastewater Reclamation Plant | Lompoc, CA | California | Liquids | 69,290 |
| Joint Water Pollution Control Plant | Los Angeles County, CA | California | Liquids | 3,500,000 |
| Hyperion Water Reclamation Plant (HWRP) | Los Angeles, CA | California | Liquids | 4,000,000 |
| Los Banos Wastewater Treatment Plant | Los Banos, CA | California | Liquids | 42,000 |
| City of Madera, Wastewater Treatment Plant | Madera, CA | California | Solids | 67,944 |
| Mammoth Community Water District | Mammoth, CA | California | Liquids | 35,000 |
| Monterey One Water - Regional Treatment Plant | Marina, CA | California | Liquids | 262,000 |
| Merced Wastewater Treatment Plant | Merced, CA | California | Solids | 91,000 |
| Sewerage Agency of Southern Marin Wastewater Treatment Plant | Mill Valley, CA | California | Solids | 30,000 |
| Modesto's Sutter Primary Treatment Facility | Modesto, CA | California | Solids | 230,000 |
| Soscol Water Recycling Facility | Napa, CA | California | Solids | 83,300 |
| [Newark Basin] - Raymond A. Boege Alvarado WWTP | Newark, CA | California | Liquids | 47,229 |
| Novato Sanitary District | Novato, CA | California | Liquids | 53,000 |
| East Bay Municipal Utility District | Oakland, CA | California | Solids | 740,000 |

| | | | | |
|---|---|---|---|---|
| Oceanside Water Pollution Control Plant * | Oceanside, San Francisco, CA | California | Solids | 250,000 |
| Regional Water Recycling Plant No.1 (RP-1) | Ontario, CA | California | Solids | 890,000 |
| Calera Creek Water Recycling Plant | Pacifica, CA | California | Liquids | 40,000 |
| Palo Alto Regional Water Quality Control Plant * | Palo Alto, CA | California | Solids | 236,000 |
| City of Paso Robles Wastewater Treatment Plant | Paso Robles, CA | California | Solids | 31,037 |
| Ellis Creek Water Recycling Facility | Petaluma, CA | California | Liquids | 65,000 |
| Silicon Valley Clean Water * | Redwood City, CA | California | Solids | 199,000 |
| Regional Treatment Plant | Regional, Laguna Niguel, CA | California | Liquids | 129,000 |
| Riverside Water Quality Control Plant | Riverside, CA | California | Liquids | 350,000 |
| Sacramento Regional Wastewater Treatment Plant * | Sacramento, CA | California | Solids | 1,480,000 |
| E.W. Blom Point Loma Wastewater Treatment Plant | San Diego, CA | California | Liquids | 2,200,000 |
| San Jose-Santa Clara Regional Wastewater Facility * | San Jose, CA | California | Solids | 1,500,000 |
| City of San Leandro Water Pollution Control Plant | San Leandro, CA | California | Liquids | 50,000 |
| City of San Mateo & Estero M.I.D. Water Quality Control Plant | San Mateo, CA | California | Solids | 150,000 |
| Central Marin Sanitation Agency | San Rafael, CA | California | Liquids | 104,250 |
| City of Santa Cruz WTF - County Influent | Santa Cruz County, CA | California | Solids | 160,000 |
| City of Santa Cruz WTF - City Influent | Santa Cruz, CA | California | Solids | 160,000 |
| City of Santa Rosa, Laguna Treatment Plant | Santa Rosa, CA | California | Solids | 230,000 |
| Sausalito-Marin City Sanitary District | Sausalito, CA | California | Liquids | 18,000 |
| South Bay International Wastewater Treatment Plant | South San Diego, CA | California | Solids | 1,600,000 |

| | | | | |
|---|---|---|---|---|
| Southeast Waste Pollution Control Plant * | Southeast San Francisco, CA | California | Solids | 750,000 |
| City of Sunnyvale Water Pollution Control Plant * | Sunnyvale, CA | California | Solids | 153,000 |
| Turlock Regional Water Quality Control Facility | Turlock, CA | California | Liquids | 86,000 |
| [Union City Basin] - Raymond A. Boege Alvarado WWTP | Union City, CA | California | Liquids | 68,150 |
| Vallejo Flood and Wastewater District Wastewater Treatment Plant | Vallejo, CA | California | Liquids | 121,000 |
| West County Wastewater District | West Contra Costa County, CA | California | Liquids | 100,000 |
| Central Marin Sanitation Agency - West Railroad | West Railroad, San Rafael, CA | California | Liquids | 25,000 |
| Windsor Wastewater Treatment, Reclamation, and Disposal Facility | Windsor, CA | California | Liquids | 28,000 |
| Winters - East Street Pump Station | Winters, CA | California | Liquids | 7,286 |
| Woodland Water Pollution Control Facility | Woodland, CA | California | Liquids | 59,000 |
| Parker Water and Sanitation District North Water Reclamation Facility | North, Parker, CO | Colorado | Liquids | 35,000 |
| Parker Water and Sanitation District South Water Reclamation Facility | South, Parker, CO | Colorado | Liquids | 25,000 |
| Stamford Water Pollution Control Authority (WPCA) | Stamford, CT | Connecticut | Solids | 140,000 |
| Seaford Wastewater Treatment Facility | Seaford, DE | Delaware | Solids | 13,172 |
| Altamonte Springs Regional Water Reclamation Facility | Altamonte Springs, FL | Florida | Liquids | 95,000 |
| Eastern Water Reclamation Facility | Eastern, Orange County, FL | Florida | Solids | 195,299 |
| Loxahatchee River Environmental Control District | Jupiter, FL | Florida | Liquids | 90,000 |
| MDWASD Central District WWTP | Key Biscayne, FL | Florida | Liquids | 829,725 |
| MDWASD North District WWTF | North Miami, FL | Florida | Liquids | 776,150 |

| | | | | |
|---|---|---|---|---|
| Northeast Water Reclamation Facility | Northeast, Saint Petersburg, FL | Florida | Liquids | 89,847 |
| Northwest Water Reclamation Facility | Northwest, Orange County, FL | Florida | Solids | 66,690 |
| Northwest Water Reclamation Facility | Northwest, Saint Petersburg, FL | Florida | Liquids | 94,218 |
| MDWASD South District WWTF | South Miami, FL | Florida | Liquids | 920,528 |
| South Water Reclamation Facility | South, Orange County, FL | Florida | Solids | 183,009 |
| Hamlin Water Reclamation Facility | Southwest, Orange County, FL | Florida | Solids | 50,000 |
| Southwest Water Reclamation Facility | Southwest, Saint Petersburg, FL | Florida | Liquids | 47,790 |
| TPSmith Water Reclamation Facility | Tallahassee, FL | Florida | Liquids | 212,065 |
| Big Creek Water Reclamation Facility | Big Creek, Roswell, GA | Georgia | Liquids | 189,593 |
| Camp Creek Water Reclamation Facility | College Park, GA | Georgia | Liquids | 73,821 |
| South Columbus Water Resources Facility | Columbus, GA | Georgia | Solids | 278,000 |
| Johns Creek Environmental Campus | Johns Creek, Roswell, GA | Georgia | Liquids | 84,486 |
| Little River Water Reclamation Facility | Little River, Roswell, GA | Georgia | Liquids | 12,818 |
| RM Clayton Water Reclamation Center | RM Clayton, Atlanta, GA | Georgia | Liquids | 294,660 |
| South River Water Reclamation Center | South River, Atlanta, GA | Georgia | Liquids | 105,160 |
| Utoy Creek Water Reclamation Center | Utoy Creek, Atlanta, GA | Georgia | Liquids | 70,887 |
| Hilo Wastewater Treatment Plant | Hilo, HI | Hawaii | Liquids | 16,257 |
| Honouliuli Wastewater Treatment Plant | Honouliuli, Honolulu, HI | Hawaii | Liquids | 300,000 |
| Kailua Regional Wastewater Treatment Plant | Kailua, Honolulu, HI | Hawaii | Liquids | 90,000 |
| Sand Island Wastewater Treatment Plant | Sand Island, Honolulu, HI | Hawaii | Liquids | 390,000 |

| | | | | |
|---|---|---|---|---|
| Wahiawa Wastewater Treatment Plant | Wahiawa, Honolulu, HI | Hawaii | Liquids | 18,000 |
| Waianae Wastewater Treatment Plant | Waianae, Honolulu, HI | Hawaii | Liquids | 44,000 |
| City of Coeur d'Alene Water Resource Recovery Facility | Coeur d'Alene, ID | Idaho | Solids | 50,540 |
| Lander Street Water Renewal Facility | Lander Street, Boise, ID | Idaho | Liquids | 108,556 |
| West Boise Water Renewal Facility | West Boise, ID | Idaho | Liquids | 186,901 |
| Glenbard Wastewater Authority | Glen Ellyn, IL | Illinois | Solids | 86,000 |
| Wheaton Sanitary District | Wheaton, IL | Illinois | Solids | 63,000 |
| Dillman Road WWTP | Bloomington, IN | Indiana | Liquids | 56,090 |
| City of Carmel WWTP | Carmel, IN | Indiana | Solids | 86,000 |
| Jeffersonville Downtown WWTP | Downtown, Jeffersonville, IN | Indiana | Liquids | 25,000 |
| North Water Reclamation Facility | North, Jeffersonville, IN | Indiana | Liquids | 25,000 |
| City of South Bend Wastewater Treatment Plant | South Bend, IN | Indiana | Liquids | 130,000 |
| City of Clinton | Clinton, IA | Iowa | Solids | 29,300 |
| Coralville Wastewater Treatment Facility | Coralville, IA | Iowa | Liquids | 23,000 |
| City of Marshalltown Water Pollution Control Plant | Marshalltown, IA | Iowa | Liquids | 27,400 |
| Muscatine STP | Muscatine, IA | Iowa | Solids | 24,400 |
| Ottumwa WPCF | Ottumwa, IA | Iowa | Liquids | 25,529 |
| Municipal Wastewater Treatment Plant No. 1 (Kaw Point) | Kaw Point, Kansas City, KS | Kansas | Solids | 90,000 |
| Lawrence Kansas River Wastewater Treatment Facility | Lawrence, KS | Kansas | Solids | 80,000 |
| Kansas City Treatment Plant #20 | P20, Kansas City, KS | Kansas | Solids | 35,000 |
| Salina Wastewater Treatment Plant | Salina, KS | Kansas | Solids | 47,000 |
| Wolcott Wastewater Treatment Facility | Wolcott, Kansas City, KS | Kansas | Liquids | 15,000 |

| | | | | |
|---|---|---|---|---|
| Morris Forman Water Quality Treatment Center | Louisville, KY | Kentucky | Solids | 423,913 |
| SWBNO East Bank Wastewater Treatment Plant | New Orleans | Louisiana | Liquids | 333,400 |
| SWBNO West Bank Wastewater Treatment Plant | New Orleans | Louisiana | Liquids | 50,500 |
| City of Bangor Wastewater Treatment Plant | Bangor, ME | Maine | Solids | 40,000 |
| Brunswick Sewer District | Brunswick, ME | Maine | Liquids | 10,000 |
| Lewiston Auburn Water Pollution Control Authority | Lewiston, ME | Maine | Liquids | 60,000 |
| Portland Water District (East End Wastewater Treatment Facility) | Portland, ME | Maine | Liquids | 65,000 |
| York Sewer District | York, ME | Maine | Liquids | 10,000 |
| Hagerstown Wastewater Treatment Plant | Hagerstown, MD | Maryland | Liquids | 90,000 |
| Marlay Taylor Water Reclamation Facility | Hollywood, MD | Maryland | Liquids | 55,000 |
| Deer Island Treatment Plant | Boston, MA | Massachu setts | Solids | 2,400,000 |
| Upper Blackstone Clean Water | Millbury, MA | Massachu setts | Liquids | 250,000 |
| City of Ann Arbor Wastewater Treatment Plant | Ann Arbor, MI | Michigan | Liquids | 125,000 |
| Jackson Wastewater Treatment Plant | Jackson, MI | Michigan | Solids | 90,000 |
| Grandville Clean Water Plant | Jenison, MI | Michigan | Solids | 75,000 |
| Mt. Pleasant WRRF | Mt. Pleasant, MI | Michigan | Liquids | 21,690 |
| Traverse City Regional Waste Water Treatment Plant | Traverse City, MI | Michigan | Liquids | 30,623 |
| City of Warren Wastewater Treatment Plant | Warren, MI | Michigan | Liquids | 140,000 |
| City of Mankato Water Resource Recovery Facility (WRRF) | Mankato, MN | Minnesot a | Solids | 70,000 |

| | | | | |
|---|---|---|---|---|
| Red Wing Wastewater Treatment Facility | Red Wing, MN | Minnesota | Solids | 16,000 |
| City Of Rochester MN Water Reclamation Plant | Rochester, MN | Minnesota | Solids | 120,000 |
| St. Cloud Nutrient, Energy and Water Recovery Facility | St. Cloud, MN | Minnesota | Liquids | 120,000 |
| 7 C- Pascagoula Moss Point POTW (Vancleave, MS) | Jackson County, MS | Mississippi | Liquids | 34,333 |
| 2C-Gautier POTW (Vancleave, MS) | Jackson County, MS | Mississippi | Liquids | 19,008 |
| Northeast Water Resource Recovery Facility | Northeast, Lincoln, NE | Nebraska | Liquids | 60,000 |
| Theresa Street Water Resource Recovery Facility | Theresa Street, Lincoln, NE | Nebraska | Liquids | 240,000 |
| Clark County Water Reclamation District (CCWRD) Flamingo Water Resource Center (FWRC) | Las Vegas, NV | Nevada | Liquids | 990,000 |
| City of Dover Wastewater Treatment Facility | Dover, NH | New Hampshire | Liquids | 30,000 |
| Hall Street Wastewater Treatment Plant | Hall Street, Concord, NH | New Hampshire | Liquids | 45,000 |
| Penacook Wastewater Treatment Facility | Penacook, Concord, NH | New Hampshire | Liquids | 4,000 |
| South Monmouth Regional Sewerage Authority | Belmar, NJ | New Jersey | Solids | 52,672 |
| Cumberland County Utilities Authority | Bridgeton, NJ | New Jersey | Liquids | 50,000 |
| The Somerset Raritan Valley Sewerage Authority | Bridgewater, NJ | New Jersey | Liquids | 130,000 |
| Passaic Valley Sewerage Commission | Newark, NJ | New Jersey | Solids | 1,500,000 |
| Township of Ocean Sewerage Authority | Oakhurst, NJ | New Jersey | Liquids | 50,000 |

| | | | | |
|---|---|---|---|---|
| Bayshore Regional Sewerage Authority | Union Beach, NJ | New Jersey | Solids | 100,000 |
| Ithaca Area Wastewater Treatment Facility | Ithaca, NY | New York | Liquids | 90,000 |
| City of Oswego Wastewater Treatment Plant | Oswego, NY | New York | Liquids | 30,000 |
| Johnnie Mosley Regional Water Reclamation Facility | Kinston, NC | North Carolina | Liquids | 25,000 |
| City of Wilson - Hominy Creek Water Reclamation Facility | Wilson, NC | North Carolina | Solids | 50,000 |
| Archie Elledge WWTP | Winston-Salem, NC | North Carolina | Liquids | 92,000 |
| Akron Water Reclamation Facility | Akron, OH | Ohio | Solids | 365,000 |
| City of Youngstown Wastewater Treatment Plant | Youngstown, OH | Ohio | Solids | 174,000 |
| DELCORA Western Regional Treatment Plant | Chester, PA | Pennsylvania | Liquids | 220,000 |
| Capital Region Water AWTF | Harrisburg, PA | Pennsylvania | Liquids | 125,000 |
| Penn State Water Treatment Facility | Penn State University Park | Pennsylvania | Liquids | 16,000 |
| City of Yankton Wastewater Treatment Facility | Yankton, SD | South Dakota | Liquids | 20,000 |
| Moccasin Bend WWTP | Chattanooga, TN | Tennessee | Liquids | 400,000 |
| M.C. Stiles Wastewater Treatment Facility | Memphis, TN | Tennessee | Liquids | 300,000 |
| DCWT Dallas | Dallas Central, Dallas, TX | Texas | Liquids | 270,000 |
| City of Gainesville Wastewater Treatment Plant | Gainesville, TX | Texas | Liquids | 17,300 |
| City of Garland Rowlett Creek WWTP | Garland, TX | Texas | Solids | 200,000 |
| Hollywood Road WWTP | Hollywood Road, Amarillo, TX | Texas | Liquids | 60,000 |
| River Road WWTP | River Road, Amarillo, TX | Texas | Liquids | 140,000 |
| South Laredo WWTP | South, Laredo, TX | Texas | Liquids | 120,000 |

| | | | | |
|---|---|---|---|---|
| Southside Wastewater Treatment Plant (City of Dallas) | Southside, Dallas, TX | Texas | Liquids | 421,700 |
| Duck Creek Wastewater Treatment Plant | Sunnyvale, TX | Texas | Solids | 186,000 |
| DCWT White Rock | White Rock Central, Dallas, TX | Texas | Liquids | 630,000 |
| Wichita Falls Resource Recovery Facility | Wichita Falls, TX | Texas | Solids | 90,000 |
| SJRA WWTF No.1 | Woodlands SJRA WWTF No. 1, TX | Texas | Liquids | 65,000 |
| SJRA WWTF No.2 | Woodlands SJRA WWTF No. 2, TX | Texas | Liquids | 70,000 |
| SJRA WWTF No.3 | Woodlands SJRA WWTF No. 3, TX | Texas | Liquids | 15,000 |
| Zacate Creek WWTP | Zacate Creek, Laredo, TX | Texas | Liquids | 140,000 |
| Central Valley Water Reclamation Facility | Central Salt Lake Valley, UT | Utah | Solids | 600,000 |
| Provo City Water Reclamation Facility | Provo, UT | Utah | Solids | 115,000 |
| City of Essex Junction Wastewater Treatment Facility | Essex Junction, VT | Vermont | Solids | 30,000 |
| Montpelier Water Resource Recovery Facility | Montpelier, VT | Vermont | Solids | 10,100 |
| South Burlington-Airport Parkway WWTF | South Burlington, VT | Vermont | Liquids | 16,000 |
| Aquia Wastewater Treatment Facility | Aquia, Stafford, VA | Virginia | Solids | 100,000 |
| Town of Hillsville Wastewater Treatment Plant | Hillsville, VA | Virginia | Solids | 3,000 |
| Little Falls Run Wastewater Treatment Facility | Little Falls Run, Stafford, VA | Virginia | Solids | 50,000 |
| City of Snohomish WWTP | Snohomish, WA | Washington | Liquids | 10,150 |
| City of Wheeling, Water Pollution Control Division | Wheeling, WV | West Virginia | Liquids | 100,000 |

| | | | | |
|---|---|---|---|---|
| Wausau Waterworks Wastewater Treatment Facility | Wausau, WI | Wisconsin | Solids | 44,000 |
| Blue Plains Advanced Wastewater Treatment Tlant | Washington DC | District of Columbia | Liquids | 2,000,000 |

**Table S2.** Other assays multiplexed with *C. auris* over the course of the study. HAV is hepatitis A virus, EVD68 is enterovirus D68, MPXV is mpox virus, HAdV_F is human adenovirus group F, HMPV is human metapneumovirus, Rota is rotavirus, HPIV is human parainfluenza virus. Dates are in month/day/year format. The fluorescent molecular is indicated after the name of the assay. FAM, 6-fluorescein amidite; HEX, hexachloro-fluorescein; Cy5, Cyanine-5; ROX, carboxyrhodamine. SCAN sites include the following wastewater treatment plants: South County Regional Wastewater Authority, City of Sunnyvale Water Pollution Control Plant, Palo Alto Regional Water Quality Control Plant, San Jose-Santa Clara Regional Wastewater Facility, Silicon Valley Clean Water, Oceanside Water Pollution Control Plant, Southeast Waste Pollution Control Plant, and Sacramento Regional Wastewater Treatment Plant, California. They are marked with an asterisk (*) In Table S1. Dates are in month/day/year format.

| | | |
|---|---|---|
| WWTPs A<br>(all other WWSCAN sites) | **9/11/23 - 10/1/23**<br><br>HAV (FAM)<br>C. auris (HEX)<br>EVD68 (FAM/HEX)<br>MPXV (Cy5)<br>HAdV_F (ROX)<br>HMPV (ATTO590)<br>Rota (ROX/ATTO590) | **10/2/23- 3/1/24**<br><br>HMPV (FAM)<br>C. auris (HEX)<br>EVD68 (FAM/HEX)<br>MPXV G2R_WA (Cy5)<br>HPIV (ROX/ATTO590) |
| WWTPs B<br>(SCAN sites) | **9/11/23 - 3/1/24**<br><br>HMPV (FAM)<br>C. auris (HEX)<br>EVD68 (FAM/HEX)<br>MPXV G2R_WA (Cy5)<br>HPIV (ROX/ATTO590) | |

**Table S3.** Samples chosen to run two *C. auris* assays, for sequencing, and running with and without a RT (reverse transcription step). The location (city and state) and date where the samples were collected is provided.

| **Samples run using two *C. auris* assays** | **Samples chosen for sequencing** | **Samples run using RT and no RT step** |
|---|---|---|
| Bangor, ME, 11/16/23<br>Bangor, ME, 9/28/23<br>Bangor, ME, 9/14/23<br>Bangor, ME, 1/8/24<br>Oakhurst, NJ, 9/26/23<br>Oakhurst, NJ, 1/4/24<br>Las Vegas, NV, 12/6/23<br>Las Vegas, NV, 12/8/23<br>Las Vegas, NV, 1/3/24 | Bangor, ME, 11/16/23<br>Bangor, ME, 9/28/23<br>Bangor, ME, 9/14/23<br>Oakhurst, NJ 9/26/23<br>Las Vegas, NV, 12/6/23 | Bangor, ME, 11/16/23<br>Bangor, ME, 9/28/23<br>Bangor, ME, 9/14/23<br>Bangor, ME, 1/8/24<br>Oakhurst, NJ, 9/26/23<br>Oakhurst, NJ, 1/4/24<br>Las Vegas, NV, 12/6/23<br>Las Vegas, NV, 12/8/23<br>Las Vegas, NV, 1/3/24 |

**Table S4.** Forward and reverse primers, and probe sequences for detection of viral nucleic acids in this study. Primers and probes were purchased from Integrated DNA Technologies (IDT, Coralville, Iowa). All probes contained fluorescent molecules and quenchers (ZEN/3′ IBFQ). ZEN, a proprietary internal quencher from IDT; IBFQ, Iowa Black FQ.

| Target | Primer/Probe | Sequence |
|---|---|---|
| BCoV | Forward | CTGGAAGTTGGTGGAGTT |
| | Reverse | ATTATCGGCCTAACATACATC |
| | Probe | CCTTCATATCTATACACATCAAGTTGTT |
| PMMoV | Forward | GAGTGGTTTGACCTTAACGTTTGA |
| | Reverse | TTGTCGGTTGCAATGCAAGT |
| | Probe | CCTACCGAAGCAAATG |
| *Candida auris, Barber et al.*[1] | Forward | CAG ACG TGA ATC ATC GAA TCT |
| | Reverse | TTT CGT GCA AGC TGT AAT TT |
| | Probe | AAT CTT CGC GGT GGC GTT GCA TTC A |

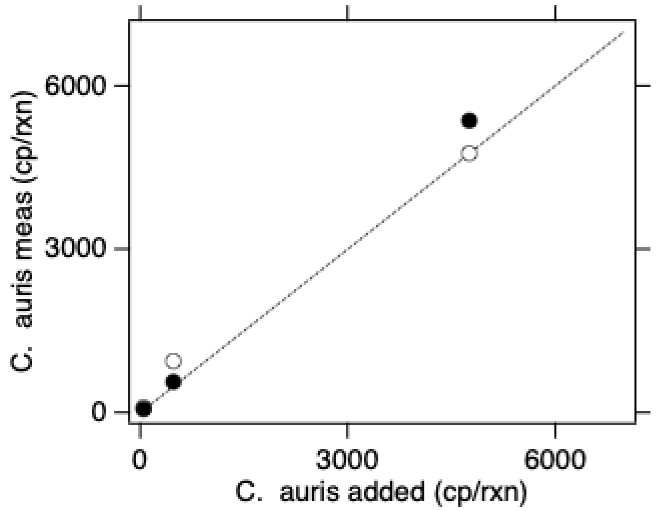


**Figure S1. Multiplex assay performance.** The concentration of the *C. auris* target in units of copies per reaction (cp/rxn) input and measured is provided. The white symbols are for reactions without the 7 background nucleic-acid targets and black symbols include high concentrations of the 7 other nucleic-acid targets. Error bars are standard deviations, if error bars cannot be seen, then they are smaller than the symbol. The line represents the 1:1 line.

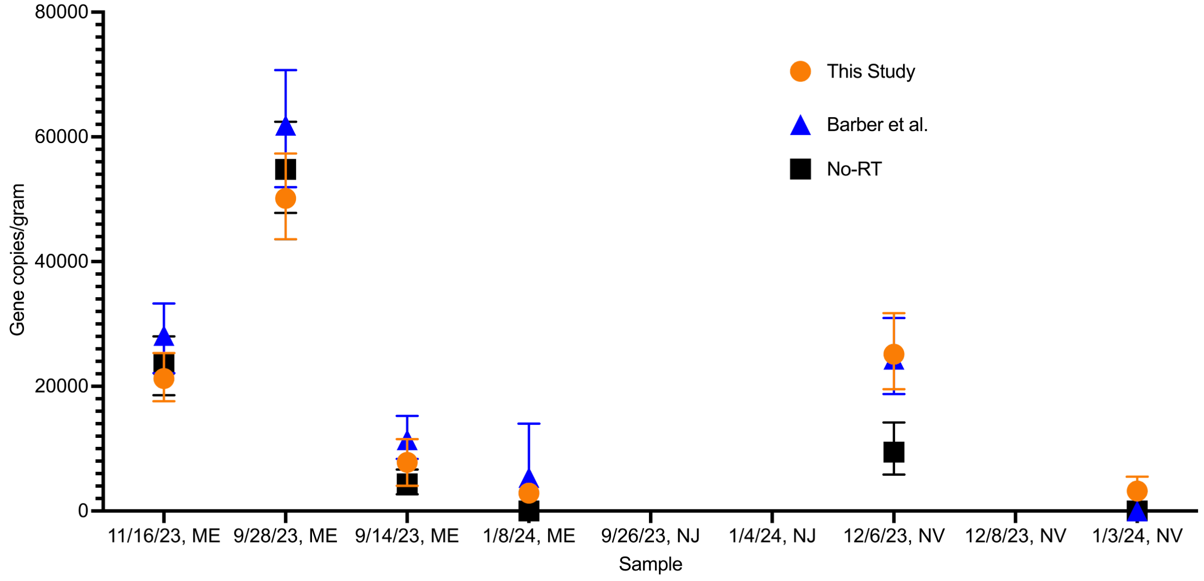


**Figure S2.** Comparison of Berber et al. assay to the assay used in this study. No significant differences were found between concentrations measured. Error bars represent standard deviations. Labels on x-axis refer to samples provided in Table S4.

| Study Description | Environmental Sampling | Sample Treatment | Sample Reduction | Nucleic-acid Extraction | Reverse Transcription | PCR Amplification | Analysis |
|---|---|---|---|---|---|---|---|
| Study name: Candida au<br>Date: March 2024<br>Completed by: A. Boehm | Notes: Decribed in methods section | Notes: No sample treatment performed | Notes: Centrifugation was used, as described in the methods | Notes: Methods provided in paper. | Notes: Performed one-step RT-PCR | Notes: droplet digital PCR used | Notes: Provided in methods |

| Control Checklist | Environmental Sampling | Sample Treatment | Sample Reduction | Nucleic-acid Extraction | Reverse Transcription | PCR Amplification | |
|---|---|---|---|---|---|---|---|
| Step performed | ☑ | ☐ | ☑ | ☑ | ☑ | ☑ | |
| Step has control info | ☐ | ☐ | ☐ | ☑ | ☑ | ☑ | Negative controls |
| # of control replicates | 0 | na | 0 | 2 | 2 | 2 | |
| Control result reported | ☐ | ☐ | ☐ | ☑ | ☑ | ☑ | |
| Method for handling failed controls described | ☐ | ☐ | ☑ | ☑ | ☑ | ☑ | |
| Step has control info | ☑ | ☐ | ☑ | ☐ | ☐ | ☐ | Positive controls |
| Control identity described | ☑ | ☐ | ☑ | ☑ | ☑ | ☑ | |
| Control quantification method described | ☑ | ☐ | ☑ | ☑ | ☑ | ☑ | |
| # control replicates | internal control | na | 6 to 10 | 6 to 10 | 6 to 10 | 6 to 10 | |
| Control result reported | ☑ | ☐ | ☑ | ☑ | ☑ | ☑ | |
| Method for handling failed controls described | ☑ | ☐ | ☑ | ☑ | ☑ | ☑ | |

### Process checklist

| Environmental Sampling | |
|---|---|
| Sample procedure | ☑ |
| Number of samples | ☑ |
| Sample amount, mean, range | ☑ |
| Sampling locations, dates, times | ☑ |
| Sample storage conditions | ☑ |
| **Sample Treatment** | |
| Treatment procedure | ☐ |
| Reagents | ☐ |
| **Sample Reduction** | |
| Reduction procedure | ☑ |
| Reagents | ☑ |
| Concentration factor | ☑ |

| Nucleic-acid Extraction | |
|---|---|
| Extraction procedure | ☑ |
| Volume or mass extracted, volume or mass obtained | ☑ |
| Extract storage conditions | ☑ |
| **Reverse Transcription** | |
| One- or two-step | ☑ |
| cDNA storage conditions (if 2 step) | ☐ |
| Reaction temperatures and times | ☑ |
| Reaction reagents and concentrations | ☑ |
| Priming method | ☑ |
| Reaction volume, added template amount | ☑ |
| RT efficiency assessment procedure (if 2-step) | ☐ |
| RT control description (if two-step) | ☐ |
| RT efficiency reported (if 2-step) | ☐ |

| qPCR or dPCR | |
|---|---|
| Target gene name, amplicon length | ☑ |
| Thermocycling temp and times | ☑ |
| Master mix composition: vendors, concentrations | ☑ |
| Additives: vendors, composition | ☑ |
| Template amount added, pre-treatment (if any) | ☑ |
| Primers: sequences, concentrations, vendors, references | ☑ |
| Amplicon confirmation method (probe, melt curve details, etc) | ☑ |
| Probe sequence, concentration, vendor, reference | ☑ |
| Instrumentation | ☑ |
| Inhibition assessment procedure | ☑ |
| Inhibition control description (if used) | ☐ |
| Number of samples tested and found inhibited | ☐ |
| Equivalent volume of sample analyzed | ☑ |

| Analysis- dPCR | |
|---|---|
| Threshold settings | ☑ |
| Technical replicates, number, well merging | ☑ |
| Partitions measured, number, mean, variance | ☑ |
| Partition volume | ☑ |
| Target copies per partition, mean, variance | ☑ |
| Program used for dPCR analysis | ☑ |
| Explanation of control results, example plots | ☑ |
| **Analysis- qPCR** | |
| Technical replicates, number, calculations | ☐ |
| Calibration standards, description, source | ☐ |
| Method of quantifying standards | ☐ |
| Calibration curve slope | ☐ |
| Calibration curve R2 | ☐ |
| Lowest standard measured or 95% LOD | ☐ |
| Cq value determination methods | ☐ |

Note to users: This checklist is provided as guidance for best practices for reporting, but is not meant to be prescriptive. Not all items in the check list will apply to all studies. Please see Borchardt et al. The Environmental

Version 3.0

This version maintained by Borchardt, Boehm, Salit, Noble, Wiggington, Spencer

**Date: 18 October 2023**

Figure S3. EMMI guidelines[2] checklist.

A

```
                1                                                                                                             112
Positive ...    TCATCGAATCTTTGAACGCACATTGCGCCTTGGGGTATTCCCCAAGGCATGCCTGTTTGAGCGTGATGTCTTCTCACCAATCTTCGCGGTGGCGTTGCATTCACAAAATTAC
Sample 1, F     ----CGAaTCTTTGAACGCACATTGCGCCTTGGGGTATTCCCCAAGGCATGCCTGTTTGAGCGTGATGTCTTCTCACCAATCTTCGCGGTGGCGTTGCATTCACAAAATTAC
Sample 2, F     -----------------------GGCGCCTTGGGGTATTCCCCAAGGCATGCCTGTTTGAGCGTGATGTCTTCTCACCAATCTGCGCGGTGGCGTTGCATTCACAAAAGCAC
Sample 3, F     -----------------GCACATTGCGCCTTGGGGTATTCCCCAAGGCATGCCTGTTTGAGCGTGATGTCTTCTCACCAATCTTCGCGGTGGCGTTGCATTCACAAAATTAC
Sample 4, F     --------TCTTTGAACGCACATTGCGCCTTGGGGTATTCCCCAAGGCATGCCTGTTTGAGCGTGATGTCTTCTCACCAATCTTCGCGGTGGCGTTGCATTCACAAAATTAC

..............................................................................................................................

                113                                                                                        204
Positive ...    AGCTTGCACGAAAAAAATCTACGCTTTTTTTTCGTTTTGTTGTCGCCTCAAATCAGGTAGGACTACCCGCTGAACTTA--------------
Sample 1, F     AGCTTGCACGAAAAAAATCTACGCTTTTTTTTCGTTTTGTTGTCGCCTCAAATCAGGTAGGACTACCCGCTGAACTTA------------AG
Sample 2, F     AGCTTGCACGAAAAAAATCTACGCTTTTTTTTCGTTTTGTTGTCGCCTCAAATCAGGTAGGACTACCCGCTGAACTTANCATCATCAATAAG
Sample 3, F     AGCTTGCACGAAAAAAATCTACGCTTTTTTTTCGTTTTGTTGTCCCCTCAAATCAGGTAATACTACCCGCTGAACTTAACNNTATCAATAAG
Sample 4, F     AGCTTGCACGAAAAAAATCTACGCTTTTTTTTCGTTTTGTTGTCGCCTCAAATCAGGTAGGACTACCCGCTGAACTTA-----------AGN

..............................................................................................................................
```

B

```
                1                                                                                                             112
Positive ...    ---------------CAGCGAAATGCGATACGTAGTATGACTTGCAGACGTGAATCATCGAATCTTTGAACGCACATTGCGCCTTGGGGTATTCCCCAAGGCATGCCTGTTT
Sample 1, R     CATCGATGAATCACGCAGCGACATGCGATACGTAGTATGACTTGCAGACGTGAATCATCGAATCTTTGAACGCACATTGCGCCTTGGGGTATTCCCCAAGGCATGCCTGTTT
Sample 2, R     GATGAATG---TACGCAGCGAAATGCGATACGTAGTATGACTTGCAGACGTGAATCATCGAATCTTTGAACGCACATTGCGCCTTGGGGTATTCCCCAAGGCATGCCTGTTT
Sample 3, R     CATCGATGAANCACGCAGCGAACTTCGATACGTAGTATGACGTGCAGACGTGAATCCNCGAATCTTCGGACGCACATTGCCCCTTGGGGTATTCCCCAAGGCATGCCTGTTT
Sample 4, R     --------------------------------TAGTATGACTTGCAGACGTGAATCATCGAATCTTTGAACGCACATTGCGCCTTGGGGTATTCCCCAAGGCATGCCTGTTT

..............................................................................................................................

                113                                                                                       203
Positive ...    GAGCGTGATGTCTTTTCGCCTATGTTCANCGGTGGCGTTGCACTCACAAAACTNCA-----------------------------------
Sample 1, R     GAGCGTGATGTCTTCTCACCAATCTTCG-CGGTGGCGTTGCA-------------------------------------------------
Sample 2, R     GAGCGTGATGTCTATCAATG-----------------------------------------------------------------------
Sample 3, R     GAGCGTAATGTCTTCTCACCAATATTCG-CGGTGGCGCTGCA-------------------------------------------------
Sample 4, R     GAGCGTGATGTCTTCTCACCAATCTTCG-CGGTGGCGTTGCATTCACAAAATTACAGCTTGCACGAAAAAAATCTACGCTTTTTTTTCGTT

..............................................................................................................................
```

**Figure S4.** Sequencing of positive samples compared to sequencing of known positive controls. Mismatches are highlighted in red, while unknown bases are yellow. Figure S2A shows the alignment of using our forward primers and the resulting sequence. The top line represents our positive control, and Samples 1-4 represent our positive samples. Figure S2B shows the same results using our negative controls.

a. California

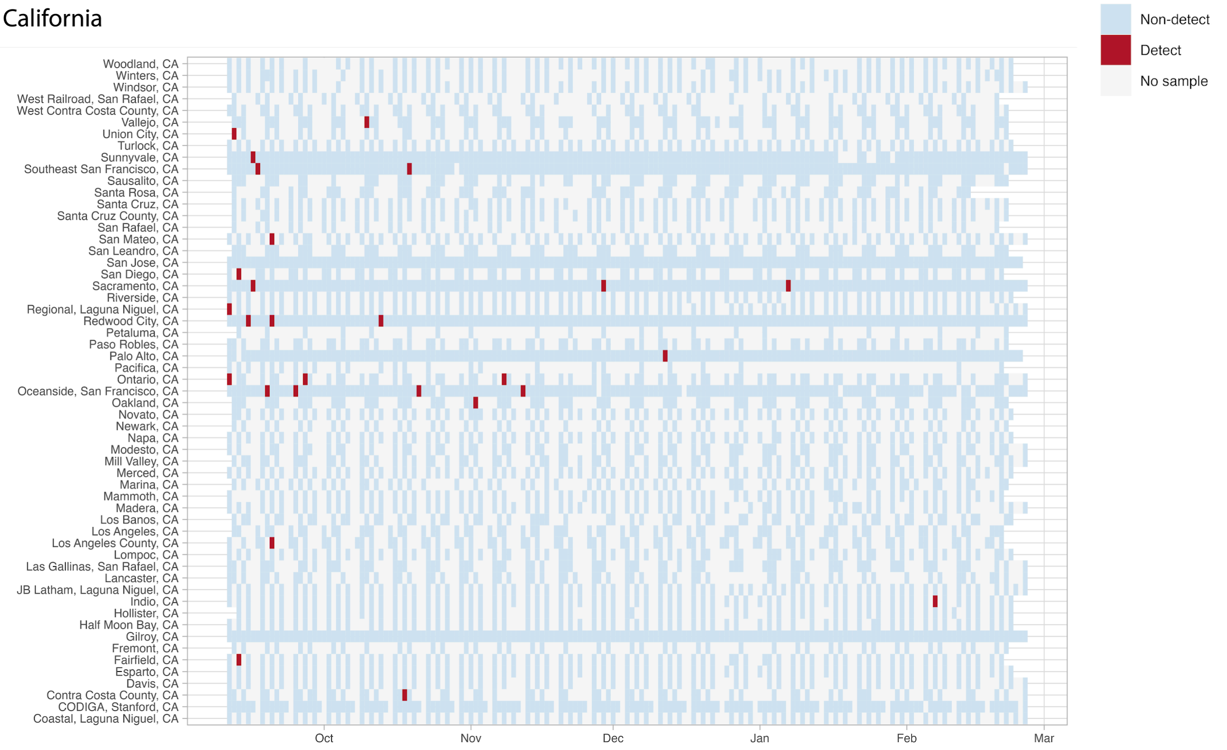


b. West

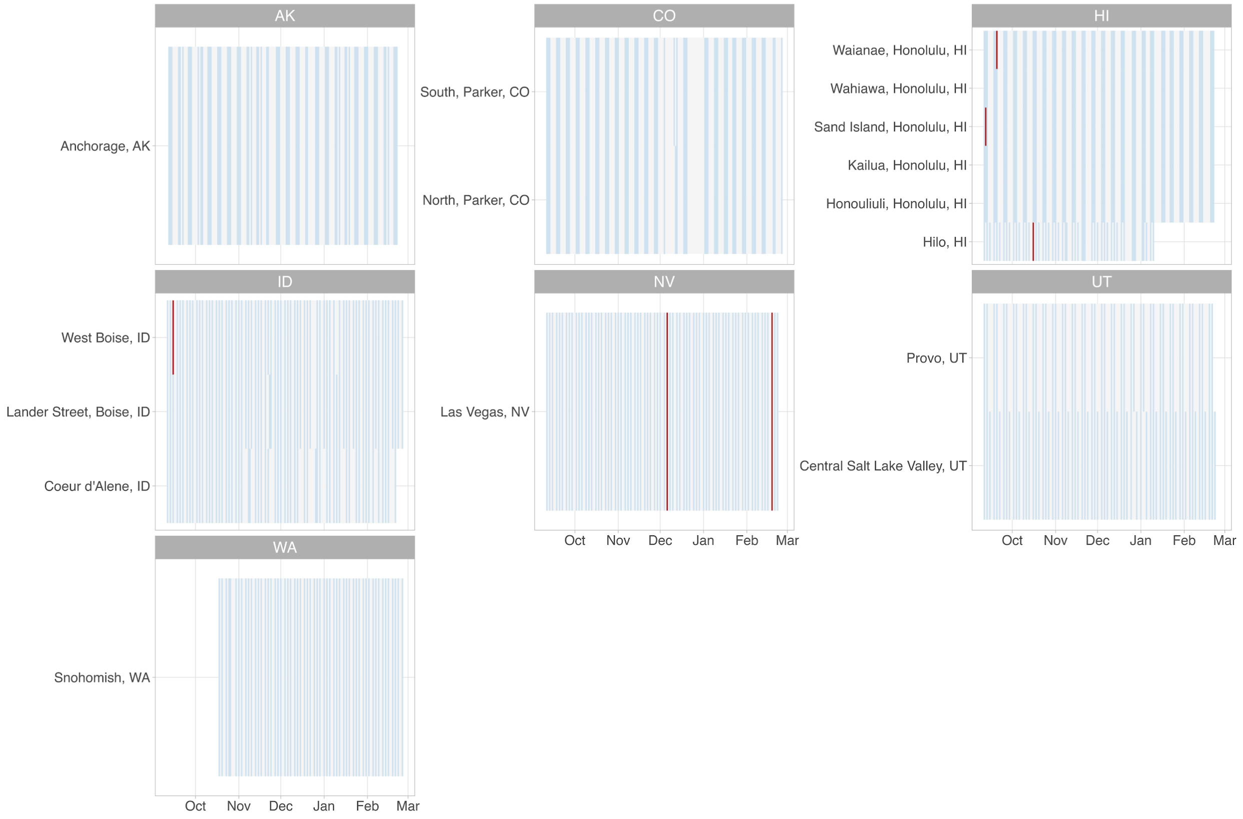

### c. South

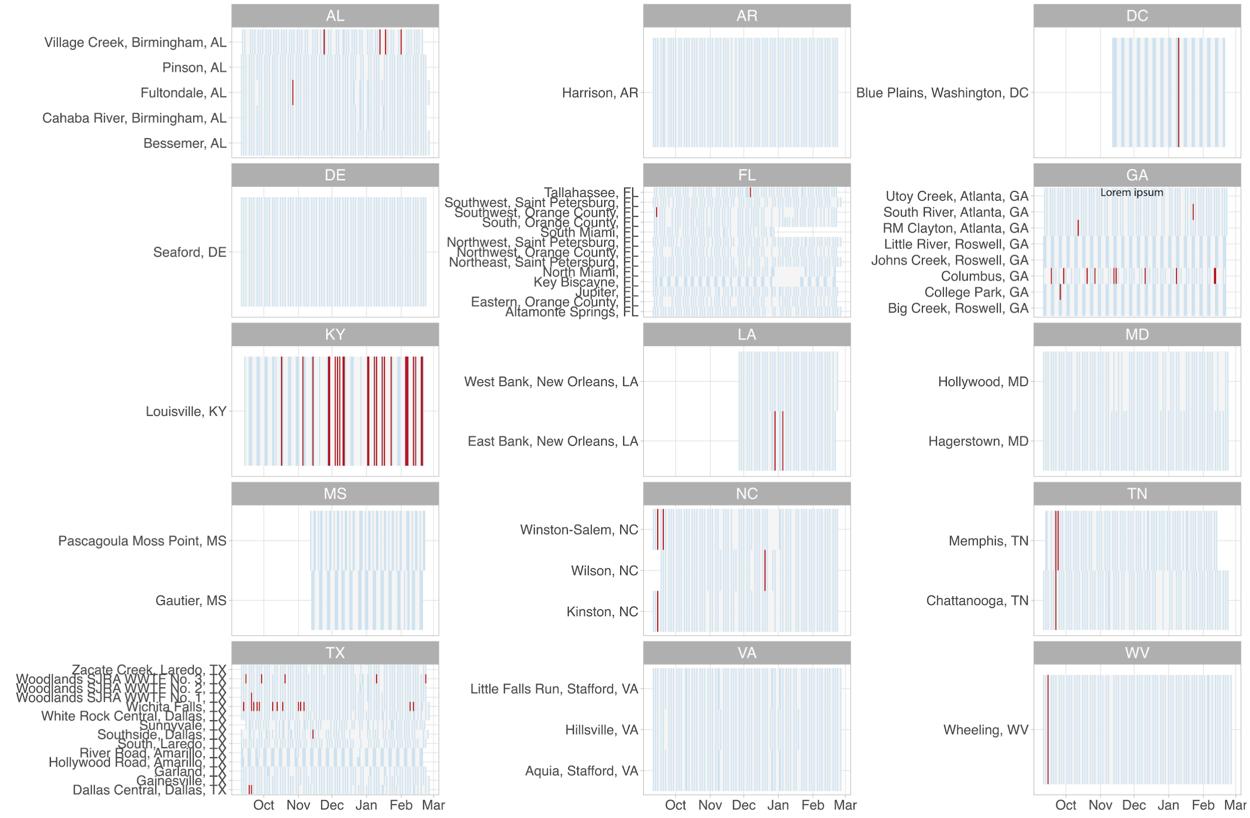

AL
Village Creek, Birmingham, AL
Pinson, AL
Fultondale, AL
Cahaba River, Birmingham, AL
Bessemer, AL
AR
Harrison, AR
DC
Blue Plains, Washington, DC
DE
Seaford, DE
FL
Tallahassee, FL
Southwest, Saint Petersburg, FL
Southwest, Orange County, FL
South, Orange County, FL
South Miami, FL
Northwest, Saint Petersburg, FL
Northwest, Orange County, FL
Northeast, Saint Petersburg, FL
North Miami, FL
Key Biscayne, FL
Jupiter, FL
Eastern, Orange County, FL
Altamonte Springs, FL
GA
Lorem ipsum
Utoy Creek, Atlanta, GA
South River, Atlanta, GA
RM Clayton, Atlanta, GA
Little River, Roswell, GA
Johns Creek, Roswell, GA
Columbus, GA
College Park, GA
Big Creek, Roswell, GA
KY
Louisville, KY
LA
West Bank, New Orleans, LA
East Bank, New Orleans, LA
MD
Hollywood, MD
Hagerstown, MD
MS
Pascagoula Moss Point, MS
Gautier, MS
NC
Winston-Salem, NC
Wilson, NC
Kinston, NC
TN
Memphis, TN
Chattanooga, TN
TX
Zacate Creek, Laredo, TX
Woodlands SJRA WWTF No. 3, TX
Woodlands SJRA WWTF No. 2, TX
Woodlands SJRA WWTF No. 1, TX
Wichita Falls, TX
White Rock Central, Dallas, TX
Sunnyvale, TX
Southside, Dallas, TX
South, Laredo, TX
River Road, Amarillo, TX
Hollywood Road, Amarillo, TX
Garland, TX
Gainesville, TX
Dallas Central, Dallas, TX
VA
Little Falls Run, Stafford, VA
Hillsville, VA
Aquia, Stafford, VA
WV
Wheeling, WV
Oct
Nov
Dec
Jan
Feb
Mar

### d. Midwest

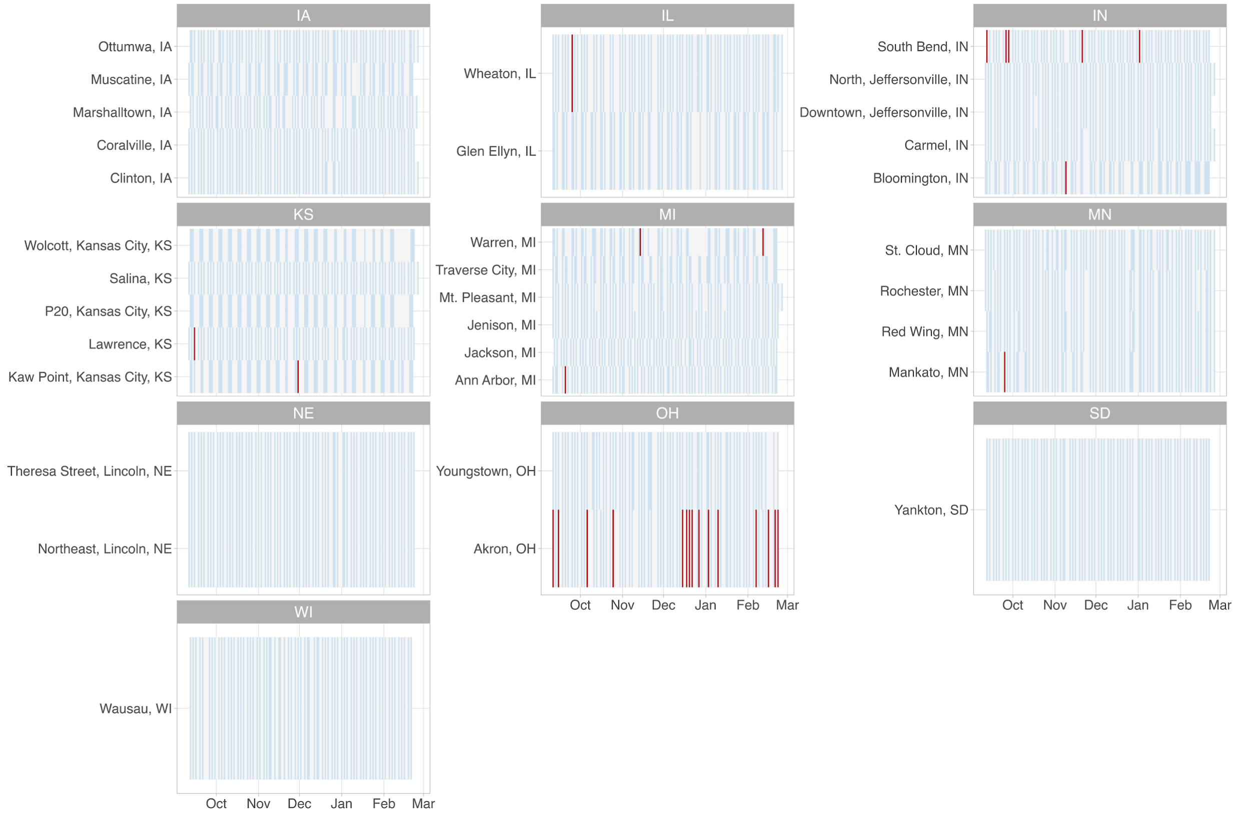
IA
Ottumwa, IA
Muscatine, IA
Marshalltown, IA
Coralville, IA
Clinton, IA
IL
Wheaton, IL
Glen Ellyn, IL
IN
South Bend, IN
North, Jeffersonville, IN
Downtown, Jeffersonville, IN
Carmel, IN
Bloomington, IN
KS
Wolcott, Kansas City, KS
Salina, KS
P20, Kansas City, KS
Lawrence, KS
Kaw Point, Kansas City, KS
MI
Warren, MI
Traverse City, MI
Mt. Pleasant, MI
Jenison, MI
Jackson, MI
Ann Arbor, MI
MN
St. Cloud, MN
Rochester, MN
Red Wing, MN
Mankato, MN
NE
Theresa Street, Lincoln, NE
Northeast, Lincoln, NE
OH
Youngstown, OH
Akron, OH
SD
Yankton, SD
WI
Wausau, WI
Oct
Nov
Dec
Jan
Feb
Mar

e. Northeast

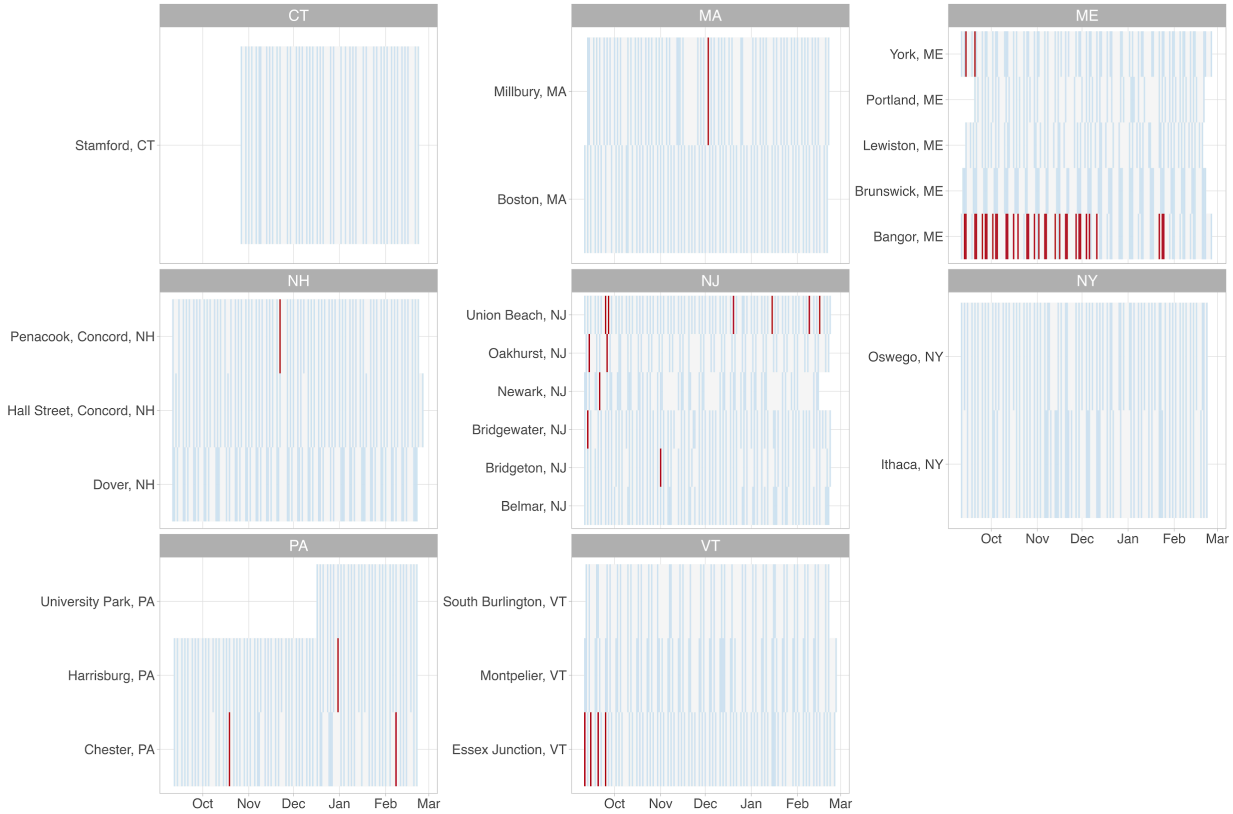


**Figure S5.** Heatmap of all national locations from September 11th, 2023 to March 1st, 2024. Graphs are organized by region and state. Figure S3a represents the locations in California, S5b is all states in the west, S5c is all states in the northeast, S5d is all states in the midwest and S5e is all states in the south.
